# Provoked cytokine response is not associated with distress or induced secondary hyperalgesia in people with suppressed HIV

**DOI:** 10.1101/2025.01.21.25320673

**Authors:** Victoria J Madden, Luyanduthando Mqadi, Gwen Arendse, Gillian J Bedwell, Ncumisa Msolo, Maia Lesosky, Mark R Hutchinson, Jonathan G Peter, Andrew Schrepf, Romy Parker, Robert R Edwards, John A Joska

## Abstract

Psychological distress predicts the onset and worsening of persistent pain, but the mechanisms that underpin this influence are poorly understood. Pro-inflammatory signalling is a plausible mechanistic link, given its known connections to distress, pain, and neural upregulation. Sustained distress may prime the inflammatory system to respond more strongly to a phasic noxious challenge, supporting neuroimmune upregulation of central nociceptive signalling and persistent pain. This cross-sectional study tested the hypotheses that *in vitro* endotoxin-provoked expression of typically pro-inflammatory cytokines (IL1β, IL6) is a partial mediator between distress and persistent pain, and that it is associated with the secondary hyperalgesia response to an experimental noxious challenge, in people with suppressed HIV. Study participants were 99 adults (mean (range) age: 43(28-64y/o; 72 females) with either no pain (n=54) or persistent pain (n=45), mostly of black South African ethnicity, low socio-economic status, and with high social support. The results replicated previous reports that distress is associated with persistent pain status and pain severity, and also showed an association between distress and the anatomical extent of pain. However, distress was not associated with provoked cytokine expression, nor was provoked cytokine expression associated with secondary hyperalgesia. The conflict between our findings and the evidence on which our hypotheses were based could reflect masking of an effect by differentially trained immune systems or a more complex relationship arising from diverse psychoneuroimmunological interactions in this sample. Our sample’s combination of HIV status, African genetic ancestry, financial impoverishment, and rich social interconnectedness is poorly represented in current research and represents an opportunity to deepen insight into psychoneuroimmunological interactions related to distress and persistent pain.

## Introduction

People with persistent pain typically report more psychological distress than their pain-free peers. Although there is some reciprocal influence between distress and pain, distress seems to influence pain more strongly than pain influences distress in both population and clinical samples; distress is a recognised risk factor for developing persistent pain [11; 17; 26; 62; 71; 83; 96]. However, despite the compelling evidence that distress influences pain, the mechanisms that support this influence are poorly elucidated.

From a psychological perspective, distress may promote pain through a bias that favours threat perception [34; 35; 50; 68; 105; 108; 116]. However, distress also alters both peripheral and spinal immunology. Sustained distress is associated with a subtle increase in tonic pro-inflammatory signalling that is thought to confer a propensity for a heightened inflammatory response —an elevated “inflammatory reactivity”—in response to a phasic challenge, such as psychosocial or physical trauma [28; 80; 99]. Experimental immune provocation, with agents such as endotoxin, models the expression of inflammatory cytokines and chemokines in response to a standardised phasic challenge, allowing insight into this inflammatory reactivity.

Inflammatory signalling has been linked to pain in both clinical samples and healthy controls. Elevated *in vitro* inflammatory reactivity (i.e. stronger inflammatory response to immune challenge) in peripheral blood is associated with clinical pain [56; 93-95] and other clinical conditions that feature pain [53; 54]. PET-detected markers of central nervous system astrocyte activation are associated with low back pain and fibromyalgia, as well as negative affect [1; 2; 64]. The extensive neuroimmune interactions within the central nervous system may explain this. In healthy controls, intravenous endotoxin exposure potentiates capsaicin-induced hyperalgesia and allodynia, indicating a priming effect of inflammation on pain, likely through heterotopic spinal facilitation of nociception [41]. Together, these data connect inflammatory signalling, nociceptive processing, and pain, but two questions remain unanswered: what influences inflammatory reactivity, and what are the consequences of inflammatory reactivity?

The current study addressed psychoneuroimmune mechanisms for persistent pain in people with suppressed HIV. HIV is a chronic infectious disease with considerable co-burdens of distress and persistent pain [23; 67; 79; 84]. In people with HIV, distress and pain seem to fluctuate together [74; 97]; distress and persistent pain are each associated with elevated expression of inflammatory signalling proteins [22; 28; 75], and inflammatory signalling is associated with neuropathic pain [98]. This study hypothesised that sustained distress primes the inflammatory system to respond more strongly to a noxious challenge (i.e. distress raises inflammatory reactivity, IR), with knock-on priming effects on central nociceptive signalling. To this end, we tested two hypotheses: (1) that IR would explain part of the association between distress and persistent pain, and (2) that IR would be positively associated with neural reactivity (i.e. stronger heterotopic facilitation in response to neural challenge, represented by induction of secondary hyperalgesia).

## Methods

### Study overview

This cross-sectional analysis uses data collected at the baseline time point (pre-designated as primary) in a longitudinal study. The data were provided by adults with HIV suppressed by antiretroviral therapy, who provided self-reported data on distress, pain, and potential confounder/collider variables, and blood samples to one assessor (‘Assessor 1’). Participants’ medical records were reviewed for clinical data. A subsample of participants also underwent experimental induction and assessment of secondary hyperalgesia (SH) on a pain-free forearm, with a second assessor (‘Assessor 2’). The Human Research Ethics Committees of the University of Cape Town and City of Cape Town (approvals 764/2019 and 24699) approved the study. The study protocol specified the two *a priori* hypotheses and the analysis approach (including planned comparisons and threshold for statistical significance) and was peer-reviewed and published before data review [69], and the study was registered at clinicaltrials.gov (NCT04757987). All deviations from the published protocol are reported in the supplementary file, and analysis scripts are available at Open Science Framework at tinyurl.com/distress-pain.

#### Blinding

Participants and Assessor 1 were blind to the study aims and hypotheses; Assessor 2 was blind to participant reports on distress levels and pain; one analyst (ML) was blinded to participant pain status for testing of Hypothesis 2 (details in supplementary file). For the SH procedure, blinding of participants and Assessor 2 was tested.

### Participants

Adults living with HIV who attended a particular primary healthcare facility in a peri-urban settlement near Cape Town, South Africa – a setting with high levels of unemployment, low income, and informal housing characteristic of poverty [101] – were recruited to participate in the study. Inclusion criteria are in Box 1.

**Box 1:** Inclusion criteria.

| Applicable to all participants: |
| --- |
| Participants $\geq 18$ and $\leq 65$ years |
| Speaks English or isiXhosa |
| HIV positive with virus suppression (*viral load* < 50 copies/ml within preceding 3 months) |
| Self-reported persistent pain (pain on most days for more than three months [7]) or no pain |
| Not pregnant (self-reported) |
| Competent to autonomously provide informed consent (assessed using explain-and-explain-back provision of study information) and able to engage in screening questionnaires |
| No neurological (self-reported) or psychiatric problems including substance abuse, psychosis, or suicide risk (screened) |
| **For subsample for SH induction:** |
| No electrical or metal implant, or current pain, in the forearm to be tested (self-report) |
| No tattoos or problems with sensation on the forearm to be tested |
| Not advised by a clinician to avoid stressful conditions (self-report) |

We aimed to recruit participants into two equally sized groups defined by pain status: one group of participants reporting no pain, the other reporting persistent pain. Individuals reporting transient or recent-onset pain were not eligible. Participants were frequency-matched across the groups on sex and age (within 5 years). Therefore, using non-probability convenience sampling, each person expressing interest in the study was screened for eligibility. Those who met the pain-related inclusion criteria were compared to the group requirements for matching. A pragmatic approach pre-determined 50 participants for each group. Consecutive eligible participants were invited to a subsample to also undergo the induction and assessment of SH, subject to additional eligibility criteria for participant safety (Box 1). Of the 50 in each group, we aimed to recruit 30 into the subsample.

### Procedure

#### Consent, blood draws, and self-report questionnaires

Using either English or isiXhosa (as chosen by the volunteer), Assessor 1 explained the study information to interested and eligible volunteers. An explain-and-explain-back approach was used to identify any cognitive difficulties and to confirm comprehension of the study procedures. After obtaining written consent, Assessor 1 used a single venepuncture to draw two blood samples and then administered the self-report questionnaires. On completion of this assessment, each participant received a ZAR150 (US$10) cash voucher. Subsample participants were given an appointment with Assessor 2 (on the same or a subsequent day).

#### Handling of blood samples

One blood sample was stimulated with endotoxin in an ex vivo closed system (TruCulture, MyriadRBM), incubated at 37 ⁰C for 24 hours, separated, and frozen at –20 ⁰C for 10-32 days, and then stored at –80 ⁰C. A second, unstimulated sample was spun and separated, and frozen for future use. Details are in the supplementary file. All stimulated samples were assayed in duplicate (R&D 3-plex Discovery assay) at a dilution factor of 1:30, using Luminex xMAP technology, to quantify the concentration of IL1β, IL6 and TNFα.

#### SH procedures for the subsample

Immediately before the SH procedure, Assessor 2 orientated and prepared the participant, obtained verbal confirmation of consent, and attached specialised electrodes (circular cathode of 10 blunt steel pins on anterior forearm; fabric anode around the upper arm). The SH procedure is shown in Figure 1. A test battery is used to assess surface area and magnitude of SH. For the post-induction test batteries only, surface area is assessed first, with a 128mN Von Frey filament (Marstock nervtest, Germany) and the 8-radial-lines method [89]. For all test batteries, five other stimulus modalities are used in a consistent order: for punctate mechanical stimuli, 128mN and 256mN blunt pinpricks (PINPRICK, MRC systems, Heidelberg, Germany); a single electrical stimulus; for dynamic light touch, a 1-second brushing stroke from a paintbrush (Prime Art Bianco Flat 16); for static light touch, a 32mN Von Frey filament. The pinprick data are used to determine the magnitude of SH; the other modalities are not reported here.

**Figure 1:**
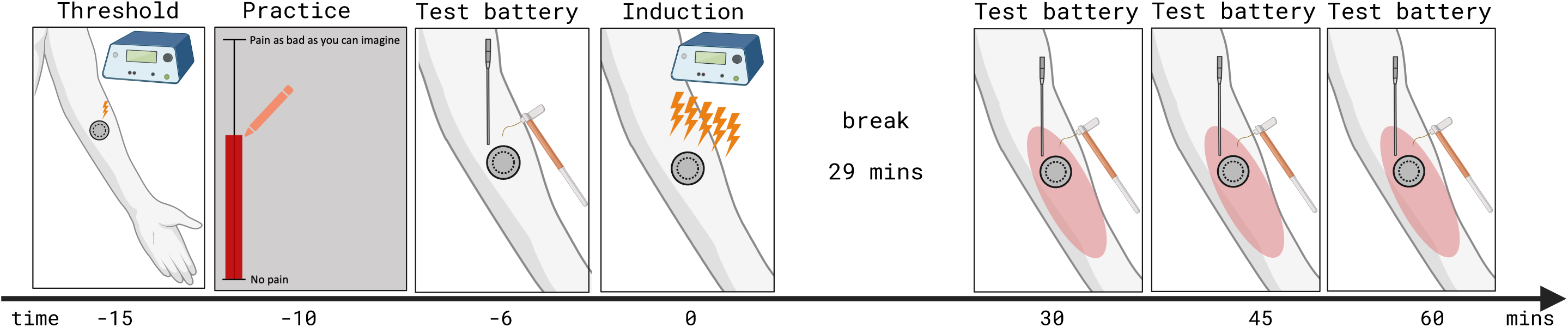
Induction and assessment of secondary hyperalgesia; not to scale. Time is shown relative to the start of the induction. Threshold identification used an adaptive staircase [6] to determine the individual electrical detection threshold. Practice: Participants practised using a touch-based visual analogue scale (VAS) to rate each stimulus in the test battery for a maximum of two batteries. Induction: Five 1-second trains of 100Hz electrical stimulation (400V, pulse width 2000μs, square pulse shape; inter-train interval 9 seconds; DS7A constant current stimulator (Digitimer Limited, Hertfordshire, UK) and Affect5 software [100] at 10 times individual detection threshold [52], with VAS rating taken for each train. Break: Participant read unrelated material. Test battery: assessment of surface area and ratings to 5 modalities. Full details in [69]. Image created in BioRender. Madden, T. (2024) https://BioRender.com/e15a892.

On completion of the SH procedure, Assessor 2 and the participant completed blinding assessments (details in supplementary file) and a brief semi-structured feedback discussion. Subsample participants were compensated with an additional ZAR150 (US$10).

### Outcomes

The main study outcomes were distress, persistent pain severity, count of persistent pain sites, provoked cytokine expression, and the surface area and magnitude of SH.

Symptoms of psychological distress were assessed with both the Hopkins 25-item Symptom Checklist (HSCL) and the World Health Organisation’s 20-item Self-Report Questionnaire (SRQ-20). The HSCL was the primary measure of distress for this study because it has been used in prior research in people living with HIV in South Africa [48; 49; 114], whereas the SRQ-20 has been used in samples from the general population of South Africa [33; 102].

Persistent pain severity and sites were assessed with an adapted version of the Brief Pain Inventory (BPI) [78]. Question sets were used to assess pain in the past week and persistent pain separately; participants provided manual ratings on the VAS and shaded the body map manually. VAS ratings were coded to lie between 0 and 10; shading of the BPI body map was coded into 18 different areas (any shading within an area constituted endorsement) and transcribed to the electronic database.

Provoked expression of IL6, IL1β, and TNFα in whole blood was used to capture IR. Of these analytes, only IL6 and IL1β had been specified *a priori*; funding unexpectedly also allowed analysis of TNFα, so we report those results here. Although antiretroviral therapy restores leukocyte counts, phenotypic alterations persist across cell types including nonclassical monocytes, neutrophils, and platelets [70; 76]. Given these changes and the continued priming of TLR4 by microbial products translocated from the gut [20], we deliberately stimulated whole blood, rather than a line of isolated cells, to capture the full functional cytokine output across functionally divergent cellular subsets. Although stimulated whole blood is traditionally considered to capture the peripheral inflammatory response, it reflects central responses well enough to provide a ‘window’ into central inflammatory responses [57].

Induced SH was assessed using both surface area (in cm^2^) and ratings. For ratings, participants rated each stimulus by swiping a stylus pen across a screen showing a vertical touch-VAS (anchors: 0 = ‘no pain’; 100 = ‘pain as bad as you can imagine’) [66]. The bar on the VAS filled up with red to the participant’s swipe point, to give visual feedback (Figure 1).

#### Adjustment variables

For Hypothesis 1, eight potential adjustment variables were assessed. Viral load, nadir CD4 count, type of antiretroviral therapy (ART), and other medications (including analgesia) were obtained from participants’ routine medical records, wherever available. When viral load results were unavailable, they were assessed as part of the study processes. Participants also self-reported medications used and known comorbidities from the three months preceding the study, including symptoms of tuberculosis and any other co-infections. The time of the blood draw was included as a covariate in all models that included IR [92; 110]. Other variables assessed were childhood trauma (28-item childhood trauma questionnaire short form, CTQ-SF), social support (Medical Outcomes Study social support survey), and stressful life events in the 12 months preceding the study (21-item Brugha Recent Life Events Scale) [69].

For Hypothesis 2, the analysis controlled for both the induction current and the VAS ratings of the five induction trains, given the current uncertainty about whether induced SH depends in part on the current used for the induction, or the subjective percept elicited by the induction procedure itself.

### Data management

Questionnaire and medical record data were captured directly into the institutional RedCap database. SH procedure data were recorded into Affect5 [100] and an Excel spreadsheet. All data were coded with a unique participant ID and stored securely. Questionnaire-based and medical data were checked for plausibility and completeness by two assessors. Errors and omissions were addressed, completed data retained, and records kept of all additions or corrections made to the data.

### Statistical analysis

#### Data handling

The de-identified data from RedCap, Affect5 and Excel were tidied and analysed in the R statistical environment version 4.3.2 (R Foundation, Vienna, Austria) [85] using RStudio version 2023.12.0+369 [104], using a private, password-protected GitHub repository. Data were visualised and described using median with 25^th^ and 75^th^ quartiles (Q1, Q3) or frequency (proportions) for numerical and categorical variables, respectively. The main R packages we used were: arsenal v. 3.6.3 [36], boot v. 1.3.28.1 [15; 19], brms v. 2.21.0 [12–14], cmdstanr v. 0.8.1 [29], epiR v. 2.0.67 [103], flextable v. 0.9.4 [103], glmmTMB v. 1.1.9 [10], lme4 v. 1.1.35.1 [4], lmeresampler v. 0.2.4 [65], lmerTest v. 3.1.3 [55], mediation v. 4.5.0 [42-46; 106], PerformanceAnalytics v. 2.0.4 [82], tidybayes v. 3.0.6 [51], and tidyverse v. 2.0.0 [113]. A full list of R packages used is provided in the analysis scripts.

Pain status was determined separately at screening and at the baseline study time point. Because some participants had a time delay between these two events, we excluded from the analysis any participant with conflicting pain status information between screening and data collection (Figure S1). To estimate cytokine levels, we fitted a weighted quadratic model to define the standard curve and used the raw fluorescence values to interpolate estimates for samples that fell outside the assay’s expected range (details in supplementary file).

#### Validity checks

To test our assumption that the HSCL would yield a reasonable estimate of psychosocial distress, we assessed its agreement with the SRQ-20 using a Bland-Altman plot and Lin’s concordance correlation coefficient (CCC) [60; 61; 111](details in supplementary file). To test whether SH was successfully induced, we constructed a mixed effects regression model with rating as the dependent variable and time of the assessment relative to the induction (coded as before/after) as the fixed independent variable. We allowed a random intercept with nesting of modality (128/256mN) within time (0, 30, 34, 60mins), and time within individual. A main effect of assessment time (indicating higher ratings after the induction than before the induction) would confirm successful induction of SH. We used zero-inflated beta regression to accommodate the high number of zeros in the data and the boundaries of the rating scale (details in supplementary file) [9; 117].

#### Planned hypothesis tests

Using data from only the persistent pain group, we first modelled the relationship between distress and each measure of persistent pain to understand this direct effect. To test whether IR mediates part of the relationship between distress and persistent pain, we conducted two cross-sectional mediation analyses for each of the 3 cytokines. We estimated the direct and indirect associations between distress (exposure) and pain severity or number of pain sites (outcome), with cytokine expression as the putative mediator.

To test whether IR was positively associated with induced SH, we used generalised mixed effects models with cytokine expression and group as independent variables. For the magnitude measure of induced SH, rating was the dependent variable. Ratings had been provided for two different stimulus weights at four different time points (before induction; 30, 45, 60 minutes after induction). The time of the assessment relative to the induction (coded as before/after) was included as a fixed independent variable. We allowed a random intercept with nesting of modality (128/256mN) within time (0, 30, 34, 60mins), and time within individual. A significant interaction between assessment time and cytokine expression would support the hypothesised association. Again, zero-inflated beta regression was used. For the surface area measure of induced SH, area was the dependent variable. The fixed independent variable of interest was the expression of the cytokine of interest, and the random intercept term specified that assessment time was nested within individual. To accommodate the high number of zeros in the data, the lack of any upper bound on the surface area of SH, and the non-parametric distribution of the data, we used a hurdle lognormal model in the ‘brms’ package, which applies Bayesian principles to modelling.

In all cases, we report both unadjusted and covariate-adjusted models, as well as effect sizes and 95% confidence intervals (CI) or credible intervals.

#### Post-hoc, exploratory analyses

To assess whether participants who withdrew had different distress severities, we used the data from all eligible participants who had commenced the SH procedure and plotted distress against procedure completed status (withdrew or completed).

## Results

### Most participants were female, low-SES, black-ethnicity South Africans

In-person assessments for the current analysis were undertaken from 11 February 2021 to 24 November 2021; the longitudinal follow-ups and medical record review continued thereafter. From the 131 volunteers who were formally screened for eligibility (see flowchart in Figure S1), data from 99 participants (72 female; median (range) age: 42 (28-64) years old) were included. There were no missing data from any of the self-reported outcomes, except three items were missing from the MOS-SSS for all participants (details in supplementary file).

Table S1 provides full demographic and economic details. All participants identified with black ethnicity; most were South African citizens (n = 98), spoke isiXhosa as their home language (n = 95), and had children (n = 97; median of 2 children each). The mean monthly income per person within each household was ZAR797 (approx. US$54). Most participants (n = 59; 60%) were unemployed, and the most common housing type was informal housing (n = 48), closely followed by own or family housing (n = 43).

A total of 47 participants (33 female; median (range) age: 41 (30-64) years old) completed the SH procedure. Five participants had withdrawn, citing the painfulness of the HFS. Data from two subsample participants were excluded for epilepsy (details in supplementary file); we report data from the remaining n = 45. One participant had been advised to avoid stressful events but enrolled on clinical judgement; their data were retained (see supplementary file). Self-report data for the subsample (n = 45) were not significantly different from the remainder (n = 54) (Table S2).

### Participants with persistent pain had higher distress, more adverse childhood experiences, moderate-severity, and multisite pain

Table 1 compares self-reported data between pain status groups; Figure 2 visualises distress and cytokine expression between groups. Psychosocial distress was higher in the persistent pain group (n = 45; median (Q1, Q3): 1.8 (1.4, 2.3)) than in the pain-free group (n = 54; 1.3 (1.1, 1.6)), Wilcoxon p < 0.001). Similarly, childhood trauma was higher in the persistent pain group (34 (28, 47)) than in the pain-free group (30 (25, 39.8)), p < 0.05). There were no between-group differences in scores for stressful life events or social support. Median (Q1, Q3) persistent pain severity score was 5.2 (4.4, 6.0) and the median number of sites of persistent pain was 3 (2, 6) (for n = 45).

**Figure 2:**
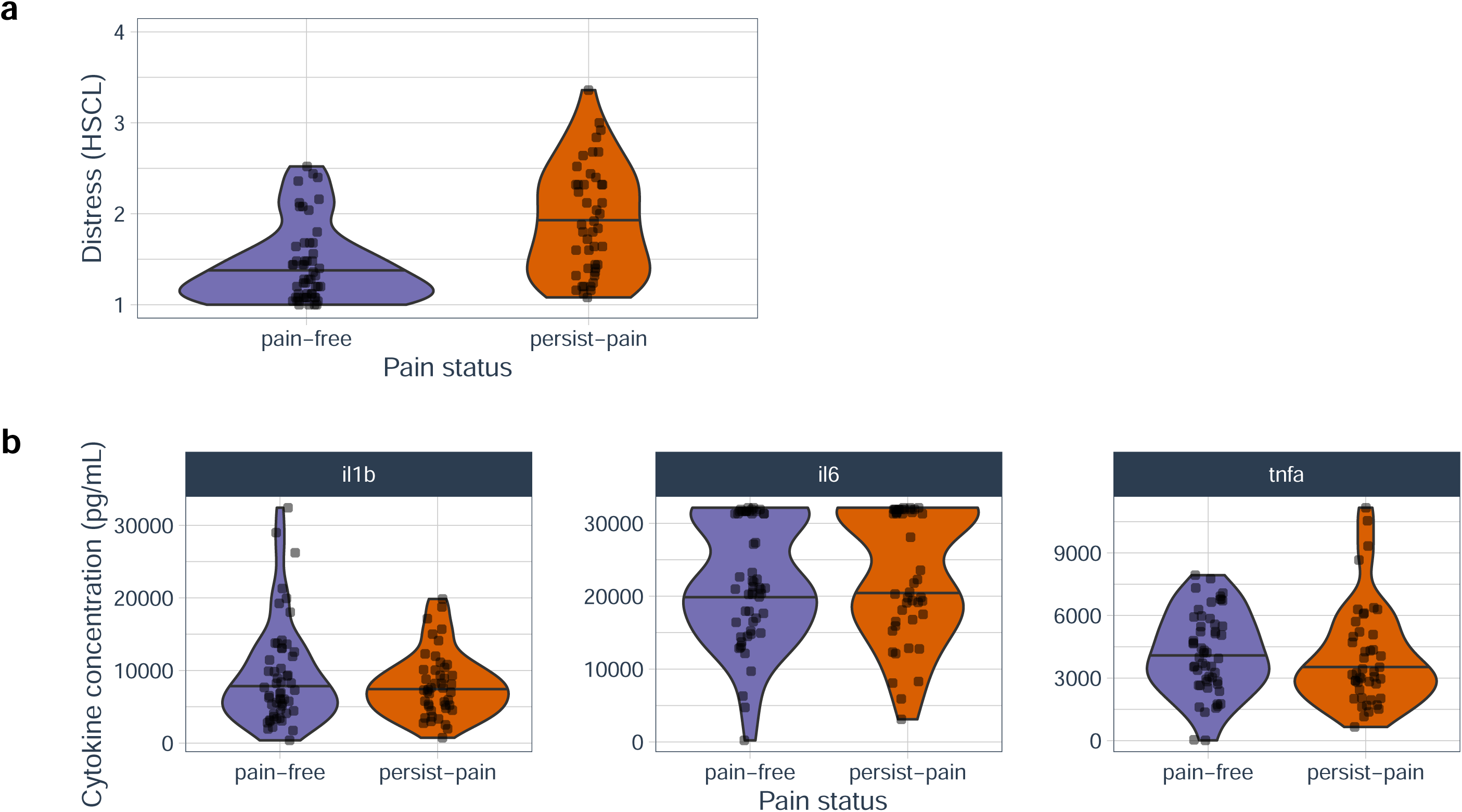
(a) Distress was lower in pain-free participants than in those with persistent pain; (b) provoked cytokine expression was no different between groups. Dots: observed data; violin shape: distribution of data; horizontal line: median.

**Table 1:**
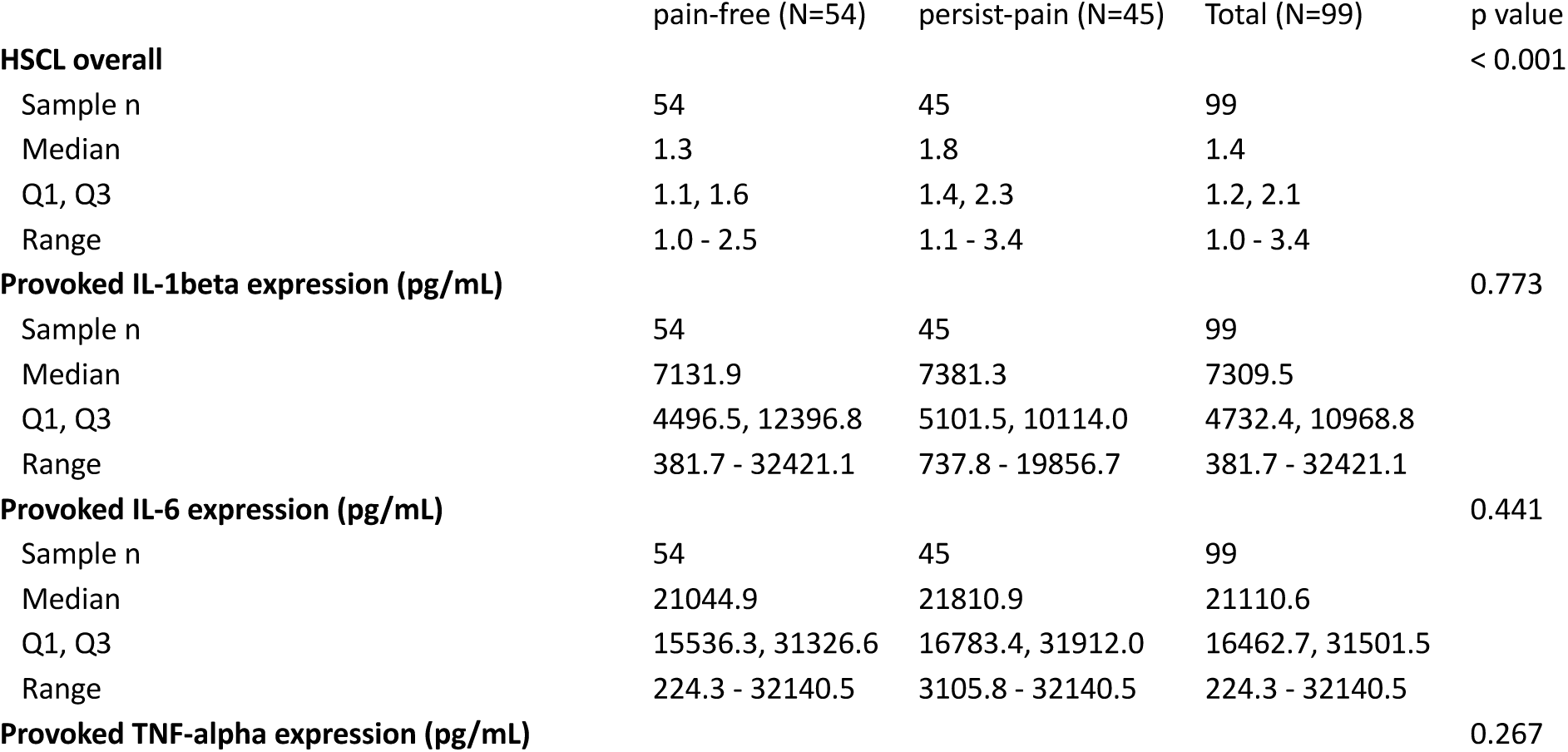
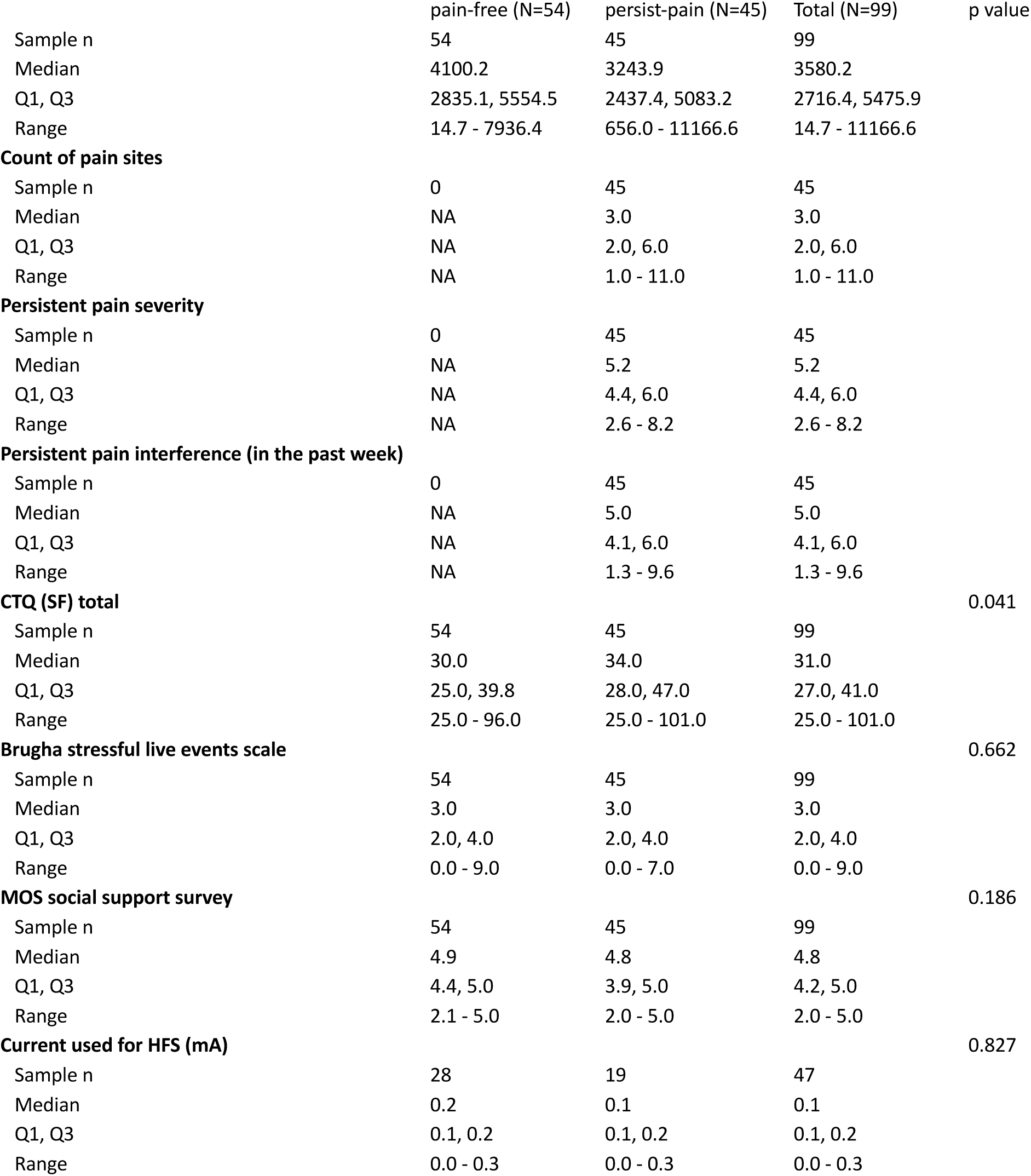
Outcomes by study group. P-values based on Wilcoxon tests. HSCL: 25-item Hopkins Symptom Checklist; CTQ (SF): Childhood Trauma Questionnaire Short Form; MOS: Medical Outcomes Survey; HFS: high-frequency electrical stimulation.

Cytokine levels fell outside the ranges of the assay for 38 (of 99) samples for IL6, 1 sample for IL1β, and 2 samples for TNFα, and were interpolated as described above. Cytokine concentration did not differ between groups (Figure 2; Table 1).

The median (IQR) current used for HFS was 0.1mA (0.1, 0.2) and was not statistically different between groups (Table 1). Ratings were missing for 119 of 225 HFS trains, due to technical error, and were imputed using the median of the available ratings for the relevant train (details in supplementary file). The group with persistent pain reported higher ratings to pinprick stimulation than the pain-free group at only the baseline time point (7.5 (0.7, 22.7) vs 1.0 (0.3, 3.4), p = 0.046) (Table S3). Neither the magnitude nor the surface area of HFS-induced SH differed between groups (Table S3). Figures S3b and S3c show pinprick ratings and SH area by time.

#### HSCL scores of distress showed good agreement with SRQ-20 scores

The Bland-Altman plot and concordance correlation coefficient showed good agreement between the estimates of distress yielded by the HSCL tool and those from the SRQ-20 tool (Figure S2 and details in supplementary file).

### Planned analyses

ART regimen, comorbidities, and other medication could not be meaningfully included as adjustment variables, because most of the sample was on first-line ART and there were too few and various comorbidities and other medications reported. The adjustment variables included in the models were: childhood trauma, stressful life events, social support, age, and sex.

#### Distress was associated with pain, but not with IR

In the 45 participants with persistent pain, distress was positively associated with both pain measures in both unadjusted and covariate-adjusted models (adjusted models shown in Figures 3a and 3b). On average, a 1-point increase in distress (on the scale with range 1-4) was associated with an increase in persistent pain severity of ∼0.8 points (unadjusted coefficient (95%CI): 0.8 (0.20-1.43), p = 0.010; adjusted 0.8 (0.18-1.47), p = 0.013) on a 0-10 scale, and two more sites of pain (unadjusted coefficient (95% CI): 2.2; (0.90-3.56), p = 0.002; adjusted 1.8 (0.16-3.42), p = 0.032) of a possible total of 18 sites. This direct association was replicated in each mediation model. Importantly however, there was no evidence to reject the null hypothesis in support of an association between distress and inflammatory reactivity (the hypothesised mediator) (Figure 3c) for any of the 3 cytokines tested. Therefore, there was no evidence that inflammatory reactivity mediated the relationship between distress and either measure of persistent pain. In the full sample of 99 participants, there was also no evidence to reject the null hypothesis in support of an association between distress and inflammatory reactivity for any of the 3 cytokines tested (Figure 3d).

**Figure 3:**
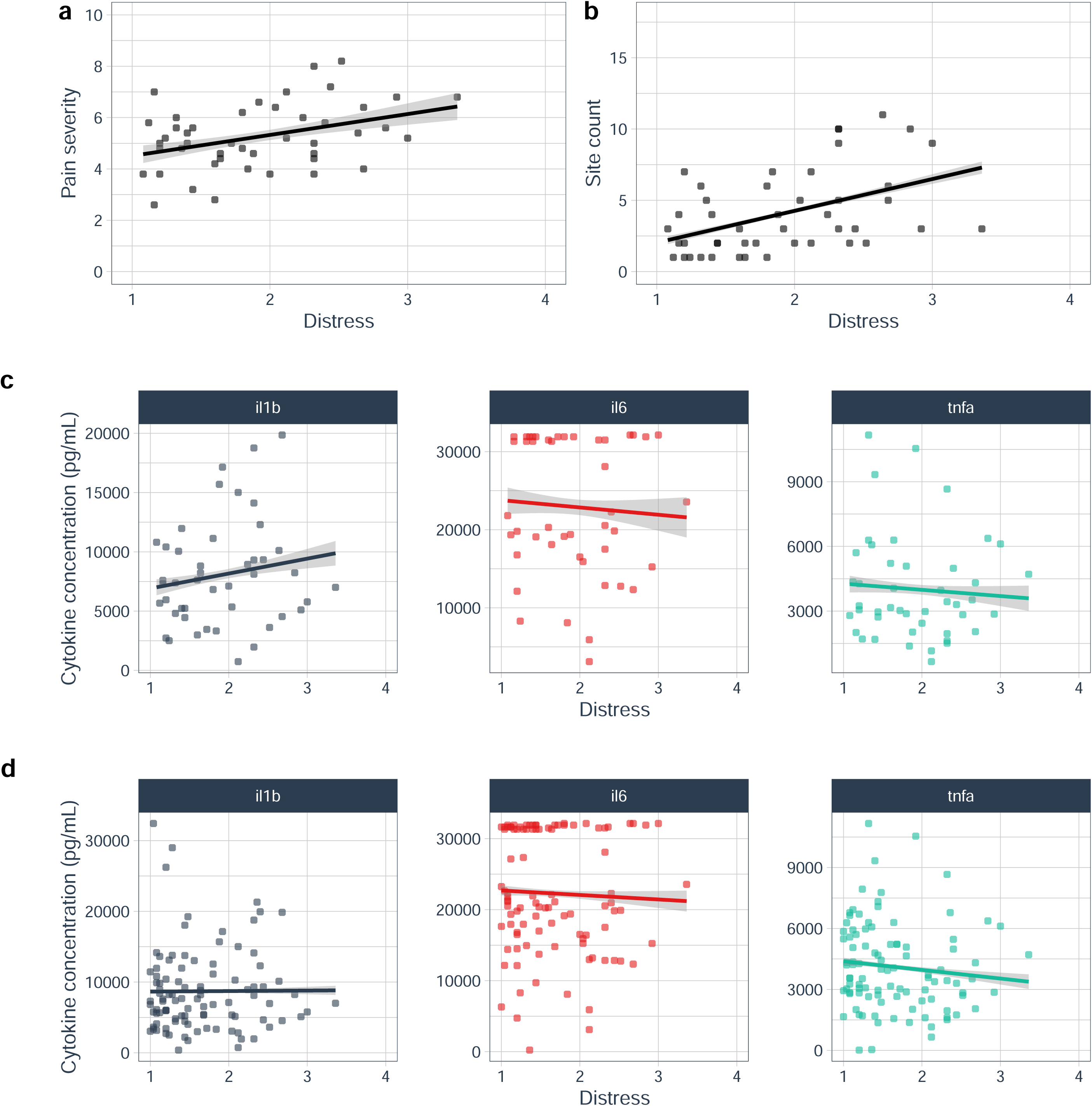
Distress was positively associated with persistent pain severity. (a) and the count of sites of persistent pain (b), but not with any of the 3 cytokines either within participants with persistent pain (c), or with all participants (d). Dots: observed data; line: predictions of adjusted linear regression model; grey ribbon: 95% CI for model-predicted values.

#### Induced secondary hyperalgesia was not associated with IR

SH was successfully induced (Figure S2): both the unadjusted and adjusted zero-inflated beta regression models showed a main effect of assessment time on rating in the zero-inflated model portions only (p < 0.001). Neither portion of the unadjusted nor adjusted regression models for the rating outcome indicated a statistically significant interaction between assessment time and cytokine expression score (Figure 4). Neither portion of the unadjusted nor adjusted hurdle models for area of SH indicated a main effect of cytokine expression score: the 95% credible intervals included zero in every case. Therefore, there was insufficient evidence to identify any association between cytokine expression score and either measure of induced SH.

**Figure 4:**
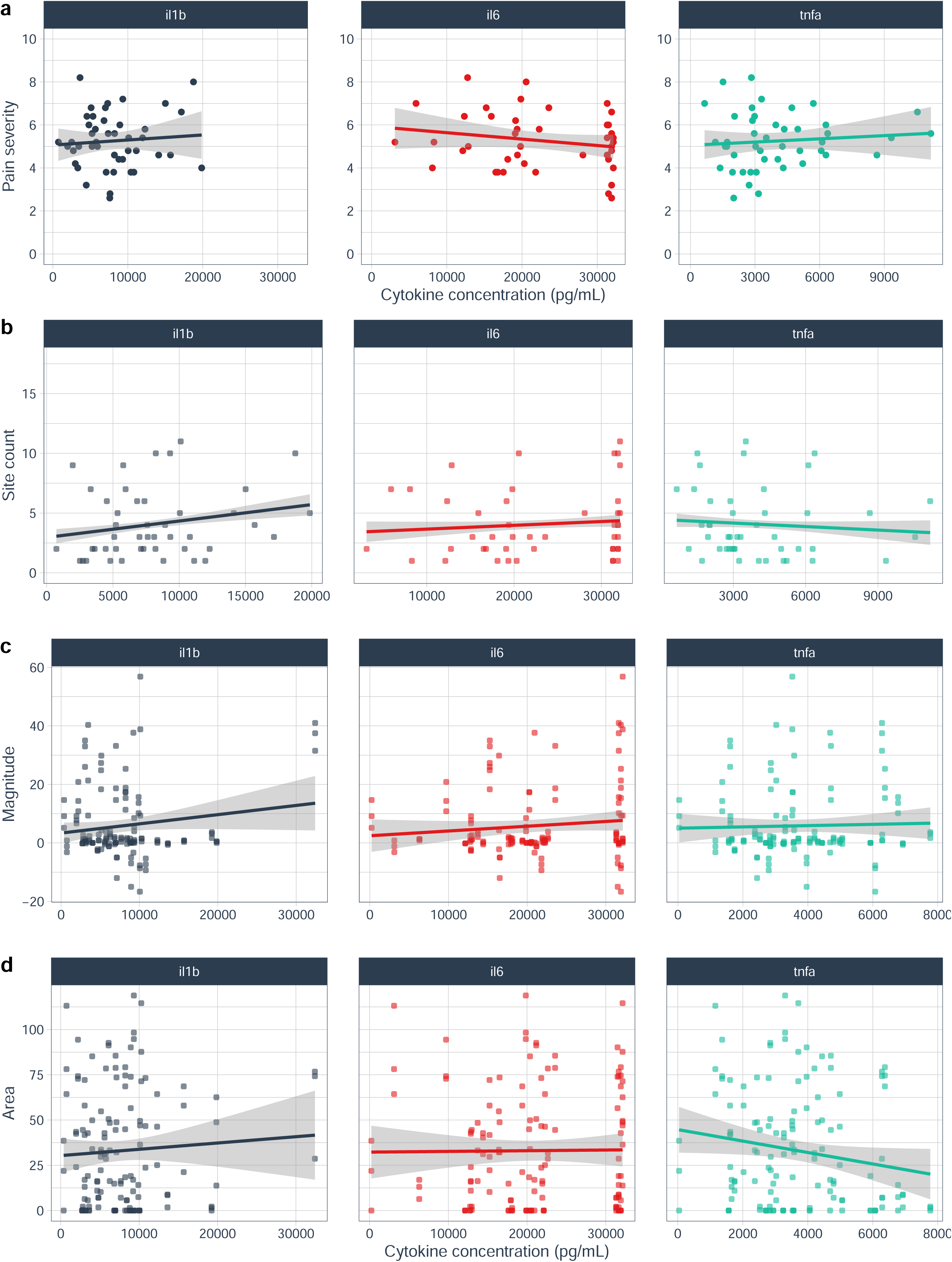
There was no evidence that cytokine expression had a linear relationship with. (a) Persistent pain severity (n=45 with persistent pain), (b) Count of painful sites (n=45 with persistent pain), (c) magnitude of SH (n=45 with SH data)(represented here with within-participant mean of baseline rating subtracted from each post-induction rating) or (d) area of SH (n=45 with SH data). Although the left panel of (b) suggests a possible association between IL1β expression and the count of sites of persistent pain, the null hypothesis was upheld. Dots: observed data; line: smoothed prediction of unadjusted linear regression; grey ribbon: 95% CI.

### Exploratory analysis

Visualisation suggested no obvious difference in distress between subsample participants who withdrew and participants who completed the procedure (Figure S4).

## Discussion

This study tested whether inflammatory reactivity partly mediates the relationship between distress and pain in people with virally suppressed HIV. We confirmed an association between distress and three measures of pain: distress was greater in participants with persistent pain than in participants with no pain, and distress was positively associated with both severity and extent of persistent pain. However, we found no evidence of an association between distress and inflammatory reactivity, as captured by endotoxin-stimulated expression of IL1β, IL6, and TNFα in whole blood. The associations between distress and both pain status and pain severity align with previous findings in the general population and a small literature in people with HIV [77; 88; 96]. In contrast to pain status and severity, few studies have focused on pain extent. In previous cross-sectional work in people with HIV in Britain and Ireland, widespread pain was more common in people with HIV than in people without HIV. The remaining prior data come from studies in the general population that have found a similar linear relationship between distress and pain extent as found in our sample of people with HIV [47; 72; 107]. That distress is robustly associated with persistent pain, and that both compromise wellbeing, suggests that people presenting with one should also be assessed for the other.

The lack of evidence for an association between distress and inflammatory reactivity in our data is intriguing, and suggests that distress has a different relationship to innate immune activity in our participants, than in participants in the prior research on which we based our hypotheses. There is a marked paucity of data on pain-related psychoneuroimmune interactions in people with genetic ancestry or social and economic circumstances that reflect those of our participants. Innate immune function –including in the Toll like receptor4 cascade recruited by endotoxin—varies with genetic ancestry and environment, although the implications of this variation for pain mechanisms have not been clarified in people with Southern African genetic ancestry [39; 109]. Social and economic circumstances may also be relevant: our participants had significant financial and food insecurity, yet high perceived social support. Given that social support is already known to buffer the immune effects of childhood adversity [87], social support may also buffer the effects of distress such that, while participants continue to notice symptoms of distress, social support attenuates a lasting influence of distress on innate immune function. Future research could test this possibility in people who report high social support in the context of psychological distress.

A competing possibility is that the hypothesised relationships do indeed exist in this sample, but are ‘masked’ by other sources of variance. Variable quantities of residual HIV and its components may introduce variance by elevating inflammatory signalling despite serological viral suppression [21; 90]. ART normalises absolute CD4+ T-cell counts but leaves a residual innate-immune signature characterised by higher basal IL1β, IL6, and TNFα, and fewer CD14^+^CD16^++^ (nonclassical) monocytes [3; 76], but lower Il1β to endotoxin provocation than HIV-controls [76]. However, our results are broadly similar to a non-ART sample of US adults (Figure 5) [94], suggesting that systematic ART-related blunting of variance is an improbable explanation for our null distress-cytokine finding. Both innate and adaptive immune responses are subject to potent ‘training’ across the life course [8; 86]. High rates of latent tuberculosis, BCG vaccination, and human herpesviruses in Southern Africa [91], alongside environment-specific mycobacteria and parasitic or fungal exposures, likely shape innate immune signalling cascades in ways that may be distinct in the Southern African context. Recent data also point to a potential role for B-cell-driven mechanisms in persistent pain, which may be especially relevant in a population with such prominent infectious immune training stimuli [58].

**Figure 5:**
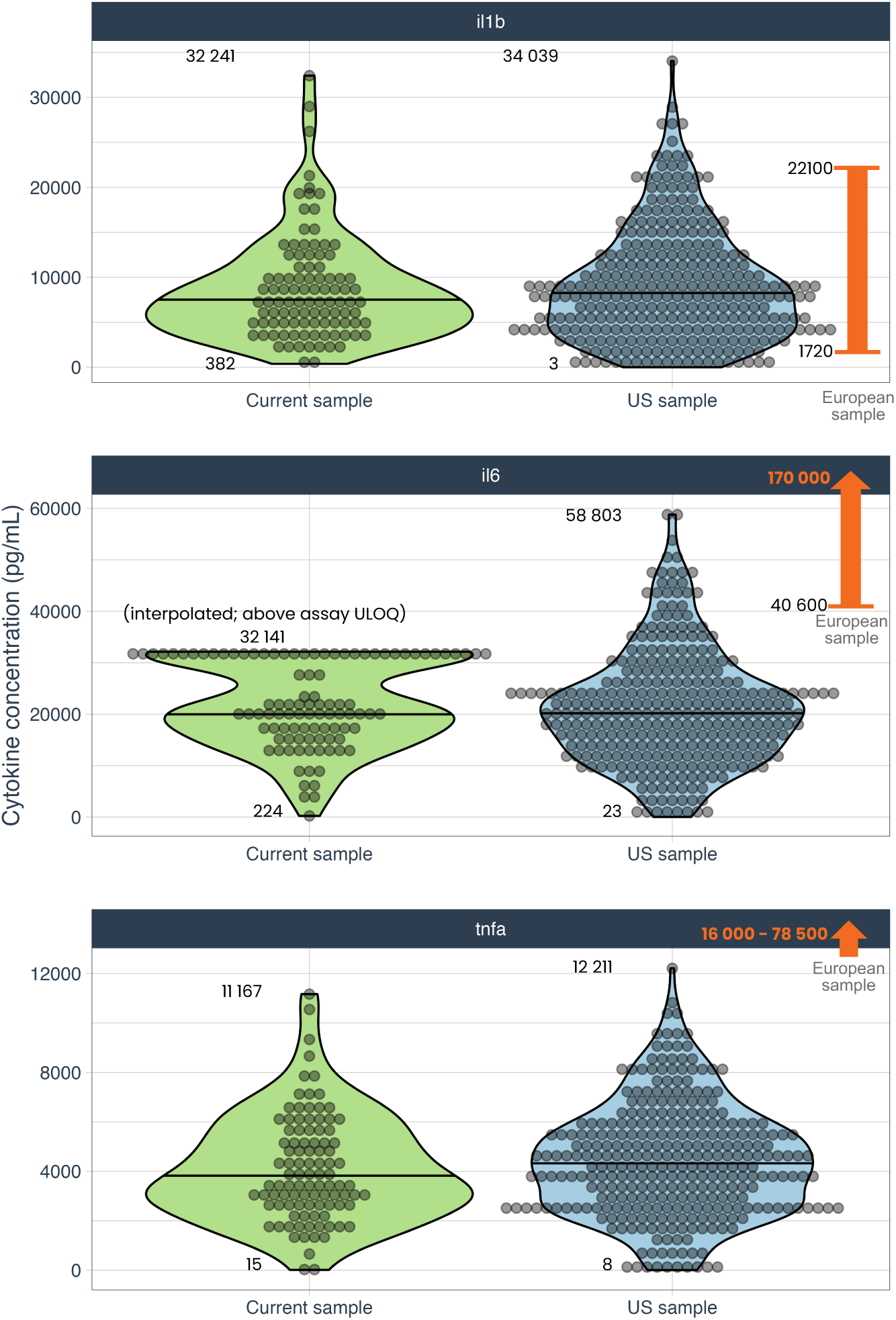
Cytokine concentrations in the current study (green) were broadly comparable to those in a study of US adults with (n= 750) and without (n=195) persistent pain (blue) [94], with the exception of IL-6, for which the assay’s upper limit of quantification (ULOQ) prevented accurate measurement in 38 of 99 samples. The results of both these studies differed notably from a European reference sample of adults (orange) who had been comprehensively immune-phenotyped as “healthy” [24]. All 3 studies used the same assay: endotoxin provocation of whole blood. Numbers in annotations indicate minima and maxima; arrows off the plot indicate numbers exceeding the axis limits.

Alternative pathways exist for the prospective relationship between general psychological distress and pain that has been shown longitudinally, elsewhere [17; 26; 62], and cross-sectionally, here. In the light of compelling evidence that pain expresses potential threat [105], it seems likely that distress is one component of a cross-system response to threat that includes other processes thought to support or exacerbate pain. Distress may either temporally precede or directly promote such processes, including enhanced transmission or reduced inhibition of nociception [30; 37; 38], hypervigilance to bodily sensations [25; 40; 59], brain-gut axis and autonomic dysregulation [27; 31], endocrine responding [16], and/or central neuroimmune activation [1; 64].

The current study’s second aim was to test the association between IR and pain-related neural reactivity in people with HIV who had either persistent clinical pain or no clinical pain. To our surprise, there was no evidence for an association between IR and induced SH, which is thought to reflect spinally mediated heterotopic facilitation [81]. One possible explanation is that concordance between inflammatory activity in the peripheral and central nervous systems is specific to conditions of neural injury or *in vivo* inflammation. Most studies correlating peripheral and central immune activity in the context of pain have used models of peripheral nerve injury [32; 57], which is known to increase immune cell migration across the blood-CNS barrier. Similarly, although experimental endotoxaemia enhances allodynia and pain sensitivity in proportion to both endotoxin dose and *in vivo* cytokine expression [41; 112], it also increases permeability of the blood-CNS barrier. In the absence of frank nerve injury or systemic inflammation, SH may be associated with central IR but not peripheral IR. In line with our previous comments on lifetime training of IR, exposures that are restricted to the peripheral compartment would also reduce peripheral-central concordance. Overcoming the current shortage of methods to sample directly from the CNS would provide an opportunity to conclusively determine whether SH is associated with central IR. A second possibility is that neural reactivity is somatotopic, as suggested by imaging data showing glial upregulation that spatially reflected the individualised distributions of clinical low back pain [64]. We induced SH on a pain-free forearm to minimise risk of exacerbating clinical pain, so we would not have captured somatotopic differences in central neural reactivity.

### Future directions

The current work highlights important outstanding questions about the relationship between distress and pain. Distress is clearly robustly associated with pain, but the nature of that association requires clarity. Achieving granular clarity will likely require at least two strategies: carefully timed longitudinal assessments of clinical distress and pain, positioned alongside systematic experimental manipulation of each variable (and potential mechanisms) to tease apart this relationship and identify and assess other factors that may confound, adjust, or strengthen it [18].

This work also highlights important missing pieces in the puzzle of HIV-associated pain: the roles of inflammatory signalling and neural reactivity, and the potential modulating roles of social context and lifetime training of immune responses. Outside the particular case of neuropathic pain, few studies have directly investigated inflammatory signalling related to HIV-associated pain in humans [22; 63; 75; 115], and none was conducted in sub-Saharan Africa, which homes the vast majority of people with HIV. Given the powerful biopsychosocial influence of environmental context, future research on pain should use a cross-system perspective to study processes within individuals and acknowledge the ongoing exchange between an individual and their surroundings. Focusing this research on the groups most affected by the health problem is likely to lead to the best clinical impact. In the case of HIV-associated pain, undertaking mechanistic research in sub-Saharan Africans would provide contextually informed and accurate mechanistic frameworks on which to base clinical intervention development and testing [73].

There are also opportunities to studying inflammatory signalling in relation to distress and persistent pain with a less potent immune challenge (such as vaccines, see [5]), by assessing unprovoked cytokine expression [21; 90], by capturing a broader range of cytokines (e.g. anti-inflammatory IL10), or by studying B-cell-mediated influences [58].

### Limitations

The current study was cross-sectional and unable to directly clarify directionality. In addition, our analysis controlled for several potential confounders, but we were unable to control for sleep problems although sleep is related to both distress and pain and may be involved in their relationship [88]. Our *in vitro* modelling of cytokine expression serves the pragmatic necessity of reducing variability due to factors such as diurnal rhythms, but cannot capture the real-time negative physiological controls that shape inflammatory signalling *in vivo* [86]. Finally, the proportion of samples with IL-6 levels out of the range of the assay warrants cautious interpretation of those data.

This cross-sectional study in people living with suppressed HIV found a positive association between psychological distress and three measures of persistent pain, but did not confirm an association between distress and an *in vitro* indicator of inflammatory reactivity. It also did not confirm an association between induced SH and inflammatory reactivity. The nature and mechanisms of the relationship between distress and pain remain to be clarified. Given the complex interplay between variables and the systems involved in nociception and pain, a cross-systems perspective is likely to be the most productive approach to this line of inquiry.

## Previous presentation of this work

The preliminary results were presented in a poster at the 2024 World Congress on Pain, in Amsterdam.

## Data Availability

The de-identified study data are available for selective sharing, subject to review of individual data requests, the use of secure computer platforms, formal use agreements, and compliance with the institutional human research ethics policies. Data use is limited to research purposes, on secure computer servers. The principal investigator (VJM) is the contact person for requests to share data.

https://osf.io/4bm5h/?view_only=427a3c0f8fb0450b96c954c36a1f24c3

## Acknowledgements

We thank Andiswa Gidana and Yoliswa Mtingeni for collecting data, and Andiswa Siyoko and Nomvula Mdwaba for recruiting participants. We thank Sarah Pedretti (University of Cape Town Lung Institute) for training and practical support with handling and storage of blood samples, Kessie Govender (Cape Peninsula University of Technology) for making the electrodes, Mathijs Franssen (KU Leuven) for research support with the SH procedure, and Peter Kamerman (University of the Witwatersrand) for his reflections on our statistical approach and interpretations. The de-identified study data are available for selective sharing, subject to review of individual data requests, the use of secure computer platforms, formal use agreements, and compliance with the institutional human research ethics policies. Data use is limited to research purposes, on secure computer servers. The principal investigator (VJM) is the contact person for requests to share data.

