## Supplementary information for "Provoked cytokine response is not associated with distress or induced secondary hyperalgesia in people with suppressed HIV"

### Supplementary file

#### Supplementary information on methods

##### Blinding

###### Participants and Assessor 1 were blinded to the study’s purpose

Participants were told neither the study aims and hypotheses, nor that they were categorised into groups. Blinding was assessed for the SH procedure by asking participants to guess the purpose of the study. In the review of the recorded responses, even a slight indication of unblinding was considered broken blinding. Assessor 1 was not told the study aims and hypotheses. Assessor 1’s blinding was not assessed.

###### Assessor 2 was blinded to questionnaire responses and VAS ratings

Assessor 2 had no access to participants’ responses to questionnaires, to blind her to distress levels and participant’s pain status. In a blinding assessment at the end of her testing of each participant, Assessor 2 guessed each participant’s pain status (options: pain-free; persistent pain) and rated her confidence in her guess (Likert scale: not at all confident, not confident, confident, and extremely confident). (Note the accidental omission of the ‘neutral’ option, explained in the supplementary file). Assessor 2 could not see the VAS on which participants provided ratings, so was blinded to rating.

###### Data analyst

One data analyst (ML) was blinded to participants’ pain status (binary variable: pain-free or persistent pain) by recoding pain status into ‘a’ and ‘b’, until the first phase of data analysis and interpretation had been completed for Hypothesis 2. Thereafter, informative labels were assigned to the study groups for further analysis, interpretation, and dissemination of the study results.

##### Handling of blood samples

One blood sample (the ‘stimulated’ sample) was drawn directly into a TruCulture® (MyriadRBM) closed system pre-loaded with endotoxin, which is the main component of the cell wall of gram-negative bacteria and a known toll-like receptor-4 stimulant. Filled tubes were incubated at 37 ⁰C for 24 hours; cells were separated from the supernatant using a seraplas® filter; tubes were frozen at -20 ⁰C for 10-32 days, and then stored at -80 ⁰C. All stimulated samples were assayed in duplicate (R&D 3-plex Discovery assay) at a dilution factor of 1:30, using Luminex xMAP technology, to estimate the levels of IL1β, IL6 and TNFα. Data for TNFα are not reported here. The second, ‘unstimulated’ sample was drawn into a vacutainer that stood upright for 30 minutes before centrifugation at 1800 revolutions per minute for ~15 minutes or until separated. Serum was pipetted into a cryotube and frozen at the same temperatures as the stimulated samples for future analysis.

##### Electrical detection threshold determination

Assessor 2 used an adaptive staircase approach [1] to determine each participant’s individual threshold for detecting a single electrical stimulus. The starting current was 0mA, increasing in steps of 0.10mA until first detection. The direction of change in current was then reversed, and the current decreased in steps of 0.5mA until undetected. The current was then increased in steps of 0.2mA until detected. The final detected current was deemed the electrical detection threshold and was multiplied by 10 for all subsequent electrical stimuli [3].

##### Estimates of cytokine levels

To estimate the levels of IL1β and IL6, we undertook the following calculations to obtain a standard curve and estimate the observed values for each panel in a reproducible manner. First, the background fluorescence was subtracted from the fluorescence observed to obtain the net florescence of each well. Second, we used the standards data to obtain an appropriate standard curve. To obtain an initial standard curve, we fitted a quadratic model to each standard, using expected concentrations (dependent variable) and net observed fluorescence (independent variable). We assessed the accuracy of this model by plotting observed vs expected standards data against a line showing model-predicted values, on both a linear scale and a log-log scale. Poor fit was apparent at the lower end of the range. We assessed this further by calculating the model-predicted values for each standard value, and the %CV between the model-predicted values and observed values. The %CV increased as the values dropped, showing poor model fit towards the lower end of the range.

To address this systematic bias in model fit, we defined a weight for each analyte and each standard as the inverse of the expected concentration (e.g. weightIL6 = 1/(expected-valueIL6)) and then fitted a quadratic model as before, but now with weighting that increased the influence of each data point in inverse proportion to its expected value. We assessed the accuracy of this weighted quadratic model by plotting observed vs expected data against a line showing model-predicted values, on both a linear scale and a log-log scale. The fit was better at the lower end of the range, although the %CV between the model-predicted values and observed values still increased (but less dramatically) as the values dropped. The weighted quadratic model was carried forward as the standard curve.

Third, we used our weighted quadratic model (standard curve) to predict the true values of the test samples from the observed net fluorescence values. Estimates that fell outside the bounds of the expected range for the panel were flagged as “out of range” and are reported for each analyte (see Results). Given that the fitted model suggested linearity of the data, we interpolated new values for each samples flagged as ‘out of range’ as follows:

For values above the expected range, we used the net fluorescence value at the maximum of the expected range (i.e. net fluorescence for Standard 1) plus half the difference between the highest (S1) and second-highest (S2) standards.

For values below the expected range, we used the net fluorescence values at the minimum of the expected range (i.e. predicted value for Standard 6) minus half the difference between the lowest (S6) and second-lowest (S5) standards.

We then used the weighted quadratic model to predict the values at each of these net fluorescence values. Finally, the mean of the two predicted values was calculated and then multiplied by 30, to account for the sample dilutions.

##### Validity checks

To test our assumption that the HSCL-25 would yield a reasonable estimate of psychosocial distress, we assessed its agreement with the SRQ-20 in two steps. First, we rescaled the total scores from the SRQ-20 (range 0-20) to match the range of the mean scores on the Hopkins-25 (range 1-4) and generated a Bland-Altman plot. Second, we calculated Lin’s concordance correlation coefficient (CCC) [4; 5], which assesses agreement between two tools measuring the same construct on a continuous scale. A Lin’s CCC closer to or equal to 1 (maximum CCC) reflects a near perfect agreement between two tools [8].

##### Hypothesis tests

To test whether IR was positively associated with the magnitude of induced SH, we used zero-inflated beta regression. Zero-inflated beta regression generates two separate models. The first is a conditional model that uses only the cases where the value of the outcome was not zero. Therefore, it estimates the relationships of the independent variables to the outcome, conditional upon the outcome being non-zero. The second is a ‘zero-inflation’ model that applies logistic regression to all the data, to estimate the relationships of the independent variables to the probability of a zero value in the outcome. In preparation for the zero-inflated beta regression, ratings of 100 were adjusted to 99.99, and then all ratings were divided by 100 to lie between 0 and 0.9999. The same structure of independent variables was used for both conditional and zero-inflated portions of the model.

To test whether IR was positively associated with the area of induced SH, we used a hurdle lognormal model. Similar to zero-inflated beta regression, hurdle models also consist of conditional and logistic regression portions with interpretations comparable to the those of zero-inflated beta regression. In addition, hurdle models assume a ‘structural’ cause for zero values — which, in this case, is appropriate because a surface area of zero indicates non-responsiveness to the induction. Given the lack of R-based function to efficiently apply a hurdle mixed-effects model in the frequentist paradigm, we used the ‘brms’ package {Bürkner, 2017 #6759;Bürkner, 2017 #6760}, which applies Bayesian principles to modelling and required us to specify sensible priors. Our choice of priors was grounded in plausibility [6]. We specified standard normal distributions for each independent variable and the intercept, and an exponential distribution for the standard deviation of the model – see supplementary file for details. To determine the mean and SD for each remaining prior, we first considered the plausible bounds of the values of each variable. Area (the outcome) has a minimum possible value of 0, but no theoretical maximum. However, a previous review [7] estimated the pooled average area of SH induced by HFS to be 34.2cm^2^ (95% CI 17-51cm^2^) from a total sample of n = 97 across four studies. Given the upper bound of the 95% CI of 51, we estimated an upper bound for the 99.9% CI of 62.41cm^2^, and increased that a little to use 63cm2 as our theoretical maximum surface area of SH.

Levels of IL6, IL1β, and TNFα were anticipated based on raw unpublished data from a previous sample of people without HIV. In those data, the 1st and 99th percentiles, respectively, were 23.25 pg/ml and 45 019.54 pg/ml for IL6, 12.69 pg/ml and 35 152.03 pg/ml for IL1β, and 41.27 pg/ml and 11 789.66 pg/ml for TNFα. We extended those bounds to 23 pg/ml and 46 000 pg/ml for IL6, 12 pg/ml and 36 000 pg/ml for IL1β, and 41 pg/ml and 12 000 pg/ml for TNFα - extending further from the maximum than from the minimum, given that HIV is anticipated to contribute to heightened cytokine expression.

Next, we calculated the parameters of a standard normal distribution (centred on 0) by considering the extreme case where each independent variable exclusively and positively determined the dependent variable - i.e. the maximum and minimum values of the predictor corresponded to areas of 63 and 0 respectively. For each of these theoretical, univariate relationships, we solved for beta by hand. Given that a standard normal distribution is defined by 99% of the distribution lying within 2.58 standard units of the mean, we then divided beta by 2.58 to obtain the standard deviation for the prior. For IL6, IL1β, we conducted these calculations after first dividing the minimum and maximum values by 10 000, in keeping with the scaling of the data for IL6, IL1β, and TNFα as entered into the models.

For the intercept, we considered the minimum values for IL6, Il1β, and TNFα were non-zero, resulting in minimum intercept values slightly below 0, at -23.25 pg/ml for IL6, -12.69 pg/ml for IL1β, and -41.27 pg/ml for TNFα. To allow for this, for area we specified a mean of 34.2 and a SD of 84.42, based on the data reported by Quesada et al. [7]. For the standard deviation of the model, we used the stanarm() [2] approach of an exponential distribution with rate of 1 divided by the actual standard deviation of the area in our data.

#### Supplementary information on results

#### Protocol deviations

Deviations from enrolment and data collection procedures are reported here. In all instances, training was repeated and deviations were reported as applicable.

**Enrolment to full study (Figure S1):**

1. One participant was enrolled in the baseline assessment despite being pregnant. Her data were excluded from analysis.
2. Three participants were enrolled in the study though they scored over 20 on the AUDIT screening tool. The data from these participants were retained since they were not intoxicated at the time of assessment and because the main purpose of excluding participants for substance abuse concerns was to limit loss to follow-up from the longitudinal component of the study. Having detected the oversight of the total AUDIT score, we included a RedCap field to auto-calculate each participant’s total AUDIT score, to support appropriate screening decisions.

**Enrolment to secondary hyperalgesia procedure:**

3. Two participants reported epilepsy and were enrolled in the SH procedure, though they should have been excluded. They reported no adverse effects. Their data were excluded from analysis.
4. One participant was enrolled in the SH procedure despite reporting having been instructed to avoid stressful events. The study nurse explained that she had judged this person to be stress-free and able to make sound decisions at the time of enrolment. The participant had reported no other exclusion criteria, and their data were retained. No adverse event was observed. The PI met in person with the study nurse to discuss this judgment about eligibility and to set up additional support for eligibility decisions.

**Age- and sex-matching:**

5. The study ID of one participant, who had been excluded upon arrival for the SH procedure due to bilateral forearm rash, was included in the age-sex matching document and only removed a month after data collection for the SH procedure ended. The removal of this participant from the document resulted in imperfect age-sex matching across the groups enrolled in the SH procedure.

**Sample handling:**

6. The box containing the unstimulated serum samples was dropped accidentally (and the lid cracked) when taken out of the -80 freezer on 30 June 2021. The samples were immediately returned to the cryobox and were back in the -80 freezer approximately 4 minutes later.

**Missing items from MOS-SSS:**

7. Three items from the MOS-SSS were not presented to any participants, due to a database error. These items were: “Someone to have a good time with”, “Someone to get together with for relaxation”, and “Someone who hugs you”. The overall within-participant score for the MOS-SSS is calculated as the mean of the scored items, and was calculated in this way – as a mean of the 16 items that were presented.

**Missing response option from blinding assessment:**

8. In the blinding assessment at the end of her testing of each participant, Assessor 2 guessed each participant’s pain status (options: pain-free; persistent pain) and rated her confidence in her guess. The Likert scale for the confidence rating described in the protocol had the 5 options of not at all confident, not confident, neutral, confident, and extremely confident. The ‘neutral’ option was accidentally omitted for all participants due to a coding error.

##### Participants

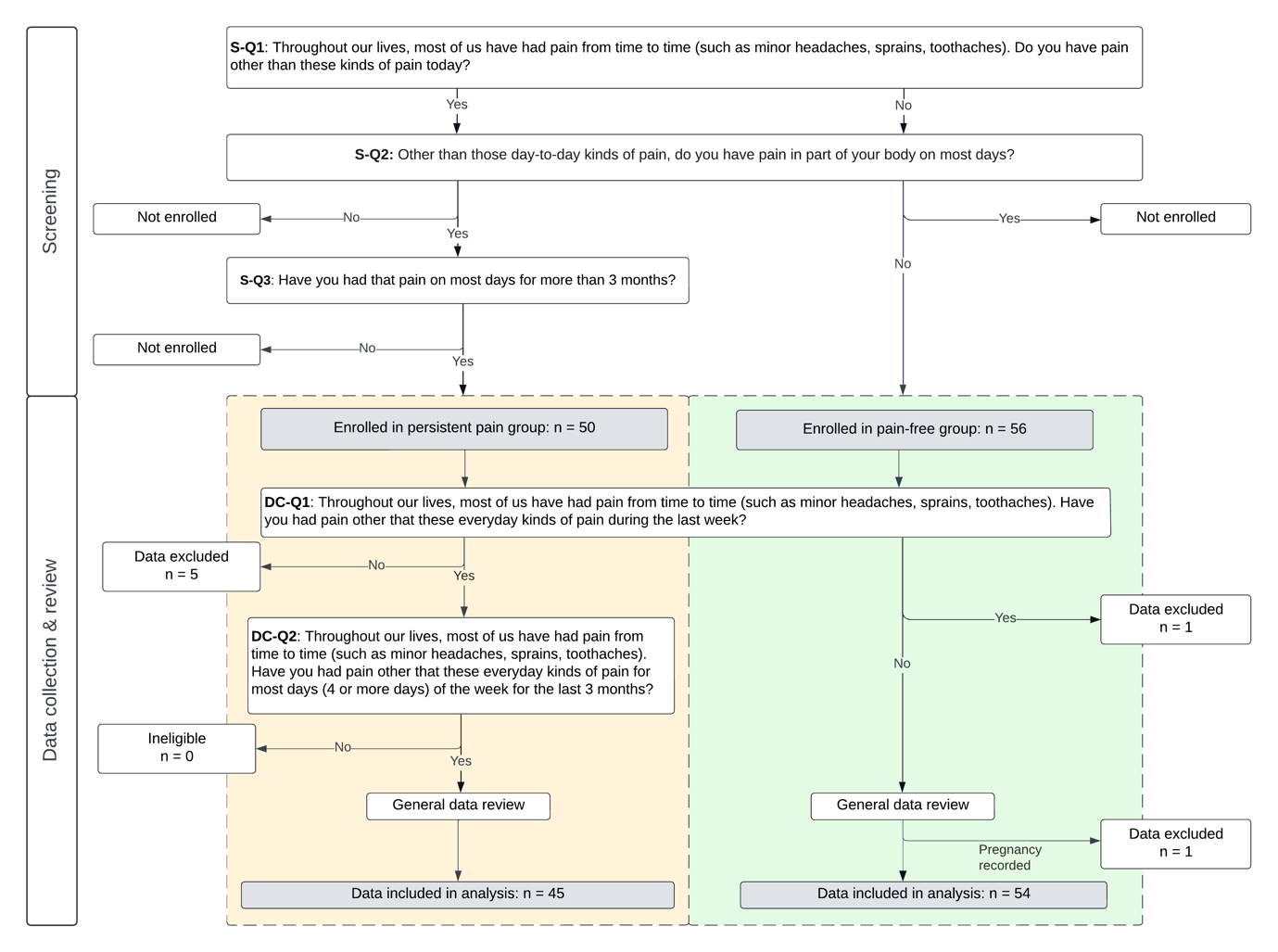

Figure S1: Screening, enrolment, and cross-checking processes to reach final sample

Table S1: Participant demographics. *Acceptable colloquial term used in specific locality of the study; typically denotes mixed ethnicity. ART: antiretroviral treatment.

|  | pain-free (N=54) | persist-pain (N=45) | Total (N=99) | p value |
| --- | --- | --- | --- | --- |
| **Sex** |  |  |  | 0.902 |
| female | 39 (72.2%) | 33 (73.3%) | 72 (72.7%) |  |
| male | 15 (27.8%) | 12 (26.7%) | 27 (27.3%) |  |
| **Age (years)** |  |  |  | 0.681 |
| Count | 54 | 45 | 99 |  |
| n missing | 0 | 0 | 0 |  |
| Median | 43 | 41 | 42 |  |
| Q1, Q3 | 37, 50 | 37, 50 | 37, 50 |  |
| Range | 28 - 64 | 28 - 63 | 28 - 64 |  |
| **Race/ethnicity** |  |  |  | 0.271 |
| black | 54 (100.0%) | 44 (97.8%) | 98 (99.0%) |  |
| black, coloured | 0 (0.0%) | 1 (2.2%) | 1 (1.0%) |  |
| **Home language** |  |  |  | 0.469 |
| afrikaans | 1 (1.9%) | 0 (0.0%) | 1 (1.0%) |  |
| afrikaans, isixhosa | 1 (1.9%) | 0 (0.0%) | 1 (1.0%) |  |
| english | 1 (1.9%) | 0 (0.0%) | 1 (1.0%) |  |
| english, afrikaans, isixhosa | 0 (0.0%) | 1 (2.2%) | 1 (1.0%) |  |
| isindebele | 0 (0.0%) | 1 (2.2%) | 1 (1.0%) |  |
| isixhosa | 49 (90.7%) | 43 (95.6%) | 92 (92.9%) |  |
| isixhosa, sesotho | 1 (1.9%) | 0 (0.0%) | 1 (1.0%) |  |
| isizulu | 1 (1.9%) | 0 (0.0%) | 1 (1.0%) |  |
| **Last educational grade/certificate passed** |  |  |  |  |
| None | 0 (0.0%) | 0 (0.0%) | 0 (0.0%) |  |
| Grade 1 | 0 (0.0%) | 0 (0.0%) | 0 (0.0%) |  |
| Grade 2 | 0 (0.0%) | 0 (0.0%) | 0 (0.0%) |  |
| Grade 3 | 0 (0.0%) | 0 (0.0%) | 0 (0.0%) |  |
| Grade 4 | 0 (0.0%) | 1 (2.2%) | 1 (1.0%) |  |
| Grade 5 | 1 (1.9%) | 0 (0.0%) | 1 (1.0%) |  |
| Grade 6 | 1 (1.9%) | 0 (0.0%) | 1 (1.0%) |  |
| Grade 7 | 1 (1.9%) | 2 (4.4%) | 3 (3.0%) |  |
| Grade 8 | 4 (7.4%) | 3 (6.7%) | 7 (7.1%) |  |
| Grade 9 | 6 (11.1%) | 7 (15.6%) | 13 (13.1%) |  |
| Grade 10 | 9 (16.7%) | 10 (22.2%) | 19 (19.2%) |  |
| Grade 11 | 15 (27.8%) | 16 (35.6%) | 31 (31.3%) |  |
| Grade 12 | 12 (22.2%) | 4 (8.9%) | 16 (16.2%) |  |
| Vocational training | 2 (3.7%) | 1 (2.2%) | 3 (3.0%) |  |
| Tertiary, university, Technikon/degree | 3 (5.6%) | 1 (2.2%) | 4 (4.0%) |  |
| Decline to answer | 0 (0.0%) | 0 (0.0%) | 0 (0.0%) |  |
| **Citizenship status** |  |  |  | 0.271 |
| Economic migrant | 0 (0.0%) | 1 (2.2%) | 1 (1.0%) |  |
| South African | 54 (100.0%) | 44 (97.8%) | 98 (99.0%) |  |
| **Religious background** |  |  |  | 0.299 |
| Christian | 42 (77.8%) | 30 (66.7%) | 72 (72.7%) |  |
| Christian, traditional | 7 (13.0%) | 12 (26.7%) | 19 (19.2%) |  |
| Muslim | 1 (1.9%) | 0 (0.0%) | 1 (1.0%) |  |
| Traditional | 4 (7.4%) | 3 (6.7%) | 7 (7.1%) |  |
| **Do you have children?** |  |  |  | 0.192 |
| No | 2 (3.7%) | 0 (0.0%) | 2 (2.0%) |  |
| Yes | 52 (96.3%) | 45 (100.0%) | 97 (98.0%) |  |
| **Number of children** |  |  |  | 0.885 |
| Count | 52 | 45 | 97 |  |
| n missing | 2 | 0 | 2 |  |
| Median | 2 | 2 | 2 |  |
| Q1, Q3 | 2, 3 | 2, 3 | 2, 3 |  |
| Range | 1 - 6 | 1 - 4 | 1 - 6 |  |
| **Marital status** |  |  |  | 0.718 |
| common-law | 12 (22.2%) | 11 (24.4%) | 23 (23.2%) |  |
| divorced | 4 (7.4%) | 4 (8.9%) | 8 (8.1%) |  |
| married | 15 (27.8%) | 7 (15.6%) | 22 (22.2%) |  |
| separated | 1 (1.9%) | 0 (0.0%) | 1 (1.0%) |  |
| single | 18 (33.3%) | 17 (37.8%) | 35 (35.4%) |  |
| single, common-law | 1 (1.9%) | 1 (2.2%) | 2 (2.0%) |  |
| widowed | 3 (5.6%) | 5 (11.1%) | 8 (8.1%) |  |
| **Monthly household income (ZAR)** |  |  |  | 0.888 |
| Count | 54 | 45 | 99 |  |
| n missing | 0 | 0 | 0 |  |
| Median | 2950 | 3000 | 3000 |  |
| Q1, Q3 | 1800, 4500 | 1650, 4300 | 1725, 4500 |  |
| Range | 0 - 17000 | 0 - 14000 | 0 - 17000 |  |
| **Monthly household income divided by people in house (ZAR)** |  |  |  | 0.785 |
| Count | 54 | 45 | 99 |  |
| n missing | 0 | 0 | 0 |  |
| Median | 900 | 714 | 797 |  |
| Q1, Q3 | 444, 1688 | 325, 1500 | 374, 1500 |  |
| Range | 0 - 3900 | 0 - 4667 | 0 - 4667 |  |
| **Social grant** |  |  |  | 0.624 |
| child | 26 (48.1%) | 17 (37.8%) | 43 (43.4%) |  |
| child, disability | 1 (1.9%) | 2 (4.4%) | 3 (3.0%) |  |
| disability | 8 (14.8%) | 7 (15.6%) | 15 (15.2%) |  |
| none | 18 (33.3%) | 18 (40.0%) | 36 (36.4%) |  |
| old age | 0 (0.0%) | 1 (2.2%) | 1 (1.0%) |  |
| old age grant | 1 (1.9%) | 0 (0.0%) | 1 (1.0%) |  |
| **Employment status** |  |  |  | 0.180 |
| casual employment | 7 (13.0%) | 9 (20.0%) | 16 (16.2%) |  |
| fulltime employment | 10 (18.5%) | 7 (15.6%) | 17 (17.2%) |  |
| retired | 0 (0.0%) | 1 (2.2%) | 1 (1.0%) |  |
| seeking employment | 0 (0.0%) | 5 (11.1%) | 5 (5.1%) |  |
| student | 1 (1.9%) | 0 (0.0%) | 1 (1.0%) |  |
| unemployed | 20 (37.0%) | 15 (33.3%) | 35 (35.4%) |  |
| unemployed, homemaker | 1 (1.9%) | 0 (0.0%) | 1 (1.0%) |  |
| unemployed, retired | 1 (1.9%) | 0 (0.0%) | 1 (1.0%) |  |
| unemployed, seeking employment | 14 (25.9%) | 8 (17.8%) | 22 (22.2%) |  |
| **Source of household income** |  |  |  | 0.793 |
| grant | 14 (25.9%) | 14 (31.1%) | 28 (28.3%) |  |
| grant, job | 1 (1.9%) | 2 (4.4%) | 3 (3.0%) |  |
| job | 33 (61.1%) | 24 (53.3%) | 57 (57.6%) |  |
| none | 6 (11.1%) | 5 (11.1%) | 11 (11.1%) |  |
| **Frequency of going without food in prev month** |  |  |  | 0.266 |
| 2-3 times | 9 (16.7%) | 9 (20.0%) | 18 (18.2%) |  |
| 4-6 times | 8 (14.8%) | 9 (20.0%) | 17 (17.2%) |  |
| More than 6 times | 8 (14.8%) | 11 (24.4%) | 19 (19.2%) |  |
| Never | 29 (53.7%) | 15 (33.3%) | 44 (44.4%) |  |
| One time | 0 (0.0%) | 1 (2.2%) | 1 (1.0%) |  |
| **Type of housing** |  |  |  | 0.210 |
| backyard housing | 2 (3.7%) | 6 (13.3%) | 8 (8.1%) |  |
| family housing | 24 (44.4%) | 19 (42.2%) | 43 (43.4%) |  |
| informal housing | 28 (51.9%) | 20 (44.4%) | 48 (48.5%) |  |
| **Living arrangement** |  |  |  | 0.431 |
| alone | 4 (7.4%) | 3 (6.7%) | 7 (7.1%) |  |
| alone, children | 0 (0.0%) | 2 (4.4%) | 2 (2.0%) |  |
| children | 15 (27.8%) | 11 (24.4%) | 26 (26.3%) |  |
| family | 2 (3.7%) | 4 (8.9%) | 6 (6.1%) |  |
| family, children | 5 (9.3%) | 7 (15.6%) | 12 (12.1%) |  |
| partner | 5 (9.3%) | 4 (8.9%) | 9 (9.1%) |  |
| partner, children | 22 (40.7%) | 12 (26.7%) | 34 (34.3%) |  |
| partner, family | 0 (0.0%) | 1 (2.2%) | 1 (1.0%) |  |
| partner, family, children | 0 (0.0%) | 1 (2.2%) | 1 (1.0%) |  |
| roommates | 1 (1.9%) | 0 (0.0%) | 1 (1.0%) |  |
| **Number of rooms in house** |  |  |  | 0.334 |
| Count | 54 | 45 | 99 |  |
| n missing | 0 | 0 | 0 |  |
| Median | 4 | 3 | 3 |  |
| Q1, Q3 | 2, 5 | 2, 5 | 2, 5 |  |
| Range | 1 - 43 | 1 - 6 | 1 - 43 |  |
| **Number of people sleeping in one room** |  |  |  | 0.170 |
| Count | 54 | 45 | 99 |  |
| n missing | 0 | 0 | 0 |  |
| Median | 2 | 2 | 2 |  |
| Q1, Q3 | 2, 3 | 2, 3 | 2, 3 |  |
| Range | 1 - 4 | 1 - 7 | 1 - 7 |  |
| **Number of people sleeping in house** |  |  |  | 0.160 |
| Count | 54 | 45 | 99 |  |
| n missing | 0 | 0 | 0 |  |
| Median | 3 | 4 | 3 |  |
| Q1, Q3 | 2, 5 | 3, 5 | 2, 5 |  |
| Range | 1 - 10 | 1 - 7 | 1 - 10 |  |
| **Facilities in house** |  |  |  | 0.534 |
| none | 0 (0.0%) | 3 (6.7%) | 3 (3.0%) |  |
| electricity | 0 (0.0%) | 1 (2.2%) | 1 (1.0%) |  |
| electricity, tv | 3 (5.6%) | 2 (4.4%) | 5 (5.1%) |  |
| electricity, tv, cd player | 0 (0.0%) | 1 (2.2%) | 1 (1.0%) |  |
| running water | 3 (5.6%) | 2 (4.4%) | 5 (5.1%) |  |
| running water, electricity | 3 (5.6%) | 2 (4.4%) | 5 (5.1%) |  |
| running water, electricity, tv | 13 (24.1%) | 10 (22.2%) | 23 (23.2%) |  |
| running water, electricity, tv, cd player | 4 (7.4%) | 1 (2.2%) | 5 (5.1%) |  |
| running water, electricity, inside toilet | 2 (3.7%) | 4 (8.9%) | 6 (6.1%) |  |
| running water, electricity, tv, inside toilet | 14 (25.9%) | 9 (20.0%) | 23 (23.2%) |  |
| running water, electricity, tv, inside toilet, cd player | 12 (22.2%) | 10 (22.2%) | 22 (22.2%) |  |
| **Do you own a cell phone?** |  |  |  | 0.454 |
| No | 1 (1.9%) | 2 (4.4%) | 3 (3.0%) |  |
| Yes | 53 (98.1%) | 43 (95.6%) | 96 (97.0%) |  |
| **How do you get to the [health] clinic?** |  |  |  | 0.710 |
| bus | 2 (3.7%) | 0 (0.0%) | 2 (2.0%) |  |
| private | 1 (1.9%) | 0 (0.0%) | 1 (1.0%) |  |
| taxi | 14 (25.9%) | 14 (31.1%) | 28 (28.3%) |  |
| walk | 34 (63.0%) | 29 (64.4%) | 63 (63.6%) |  |
| walk, taxi | 2 (3.7%) | 1 (2.2%) | 3 (3.0%) |  |
| walk, taxi, bus | 1 (1.9%) | 1 (2.2%) | 2 (2.0%) |  |
| **How much did you pay for transport to the clinic?** |  |  |  | 0.775 |
| Count | 54 | 45 | 99 |  |
| n missing | 0 | 0 | 0 |  |
| Median | 0 | 0 | 0 |  |
| Q1, Q3 | 0, 23 | 0, 12 | 0, 16 |  |
| Range | 0 - 24 | 0 - 40 | 0 - 40 |  |
| **Did you take time off work to get to the clinic?** |  |  |  | 0.226 |
| No | 48 (88.9%) | 43 (95.6%) | 91 (91.9%) |  |
| Yes | 6 (11.1%) | 2 (4.4%) | 8 (8.1%) |  |
| **Did you have to pay for someone to watch your child?** |  |  |  | 0.491 |
| I do not have a child who needs supervision | 4 (7.4%) | 1 (2.2%) | 5 (5.1%) |  |
| No | 46 (85.2%) | 41 (91.1%) | 87 (87.9%) |  |
| Yes | 4 (7.4%) | 3 (6.7%) | 7 (7.1%) |  |
| **Years since starting ART** |  |  |  | 0.178 |
| Count | 54 | 45 | 99 |  |
| n missing | 0 | 0 | 0 |  |
| Median | 9 | 8 | 8 |  |
| Q1, Q3 | 7, 12 | 5, 11 | 6, 12 |  |
| Range | 1 - 23 | 2 - 17 | 1 - 23 |  |
| **ART regimen** |  |  |  | 0.520 |
| Abacavir/Lamivudine/Atazanavir/Ritonavir | 1 (1.9%) | 0 (0.0%) | 1 (1.0%) |  |
| Abacavir/Lamivudine/Dolutegravir | 0 (0.0%) | 1 (2.2%) | 1 (1.0%) |  |
| Lamivudine/Zidovudine/Aluvia | 0 (0.0%) | 1 (2.2%) | 1 (1.0%) |  |
| Ritonavir/Lamivudine/Zidovudine/Atazanavir | 1 (1.9%) | 0 (0.0%) | 1 (1.0%) |  |
| Tenofovir/Emtricitabine/Efavirenz | 11 (20.4%) | 11 (24.4%) | 22 (22.2%) |  |
| Tenofovir/Emtricitabine/Lopinavir/Ritonavir | 0 (0.0%) | 1 (2.2%) | 1 (1.0%) |  |
| Tenofovir/Lamivudine/Dolutegravir | 38 (70.4%) | 30 (66.7%) | 68 (68.7%) |  |
| Tenofovir/Lamivudine/Efavirenz | 3 (5.6%) | 1 (2.2%) | 4 (4.0%) |  |
| **Do you hide your ART?** |  |  |  | 0.022 |
| No | 49 (90.7%) | 33 (73.3%) | 82 (82.8%) |  |
| Yes | 5 (9.3%) | 12 (26.7%) | 17 (17.2%) |  |
| **Accessed mental health care in past year** |  |  |  | 0.362 |
| None | 53 (98.1%) | 44 (97.8%) | 97 (98.0%) |  |
| Psychologist | 1 (1.9%) | 0 (0.0%) | 1 (1.0%) |  |
| Social worker | 0 (0.0%) | 1 (2.2%) | 1 (1.0%) |  |

Table S2: Comparison between data from those included and not included in subsample. P-values based on Wilcoxon tests. HSCL-25: 25-item Hopkins Symptom Checklist; CTQ (SF): Childhood Trauma Questionnaire Short Form; MOS: Medical Outcomes Survey

|  | no (N=54) | yes (N=45) | Total (N=99) | p value |
| --- | --- | --- | --- | --- |
| **Age (years)** |  |  |  | 0.578 |
| Sample n | 54 | 45 | 99 |  |
| Median | 43.0 | 41.0 | 42.0 |  |
| Q1, Q3 | 37.0, 51.0 | 35.0, 47.0 | 37.0, 50.0 |  |
| Range | 28.0 - 63.0 | 30.0 - 64.0 | 28.0 - 64.0 |  |
| **Sex** |  |  |  | 0.434 |
| female | 41 (75.9%) | 31 (68.9%) | 72 (72.7%) |  |
| male | 13 (24.1%) | 14 (31.1%) | 27 (27.3%) |  |
| **HSCL-25 overall** |  |  |  | 0.290 |
| Sample n | 54 | 45 | 99 |  |
| Median | 1.4 | 1.5 | 1.4 |  |
| Q1, Q3 | 1.2, 1.8 | 1.1, 2.2 | 1.2, 2.1 |  |
| Range | 1.0 - 3.0 | 1.0 - 3.4 | 1.0 - 3.4 |  |
| **Provoked IL-1beta expression (pg/mL)** |  |  |  | 0.211 |
| Sample n | 54 | 45 | 99 |  |
| Median | 7487.0 | 7003.8 | 7309.5 |  |
| Q1, Q3 | 5256.1, 12913.2 | 4023.5, 9349.7 | 4732.4, 10968.8 |  |
| Range | 1731.5 - 28994.9 | 381.7 - 32421.1 | 381.7 - 32421.1 |  |
| **Provoked IL-6 expression (pg/mL)** |  |  |  | 0.227 |
| Sample n | 54 | 45 | 99 |  |
| Median | 22791.1 | 20383.9 | 21110.6 |  |
| Q1, Q3 | 17118.9, 31607.2 | 15240.2, 31326.6 | 16462.7, 31501.5 |  |
| Range | 4735.9 - 32140.5 | 224.3 - 32140.5 | 224.3 - 32140.5 |  |
| **Provoked TNF-alpha expression (pg/mL)** |  |  |  | 0.115 |
| Sample n | 54 | 45 | 99 |  |
| Median | 4244.7 | 3521.6 | 3580.2 |  |
| Q1, Q3 | 2843.7, 5816.6 | 2525.8, 4710.1 | 2716.4, 5475.9 |  |
| Range | 14.7 - 11166.6 | 40.2 - 7769.8 | 14.7 - 11166.6 |  |
| **Count of pain sites** |  |  |  | 0.617 |
| Sample n | 26 | 19 | 45 |  |
| Median | 3.5 | 3.0 | 3.0 |  |
| Q1, Q3 | 2.0, 6.0 | 2.0, 4.5 | 2.0, 6.0 |  |
| Range | 1.0 - 10.0 | 1.0 - 11.0 | 1.0 - 11.0 |  |
| **Persistent pain severity** |  |  |  | 0.519 |
| Sample n | 26 | 19 | 45 |  |
| Median | 5.1 | 5.2 | 5.2 |  |
| Q1, Q3 | 4.2, 6.1 | 4.7, 6.0 | 4.4, 6.0 |  |
| Range | 2.6 - 8.0 | 3.8 - 8.2 | 2.6 - 8.2 |  |
| **Persistent pain interference (in the past week)** |  |  |  | 0.854 |
| Sample n | 26 | 19 | 45 |  |
| Median | 5.1 | 5.0 | 5.0 |  |
| Q1, Q3 | 4.2, 6.0 | 3.9, 6.1 | 4.1, 6.0 |  |
| Range | 1.3 - 9.6 | 3.0 - 9.4 | 1.3 - 9.6 |  |
| **CTQ (SF) total** |  |  |  | 0.871 |
| Sample n | 54 | 45 | 99 |  |
| Median | 30.5 | 31.0 | 31.0 |  |
| Q1, Q3 | 27.0, 40.8 | 26.0, 44.0 | 27.0, 41.0 |  |
| Range | 25.0 - 79.0 | 25.0 - 101.0 | 25.0 - 101.0 |  |
| **Brugha stressful live events scale** |  |  |  | 0.337 |
| Sample n | 54 | 45 | 99 |  |
| Median | 2.0 | 3.0 | 3.0 |  |
| Q1, Q3 | 2.0, 3.8 | 2.0, 4.0 | 2.0, 4.0 |  |
| Range | 0.0 - 9.0 | 0.0 - 7.0 | 0.0 - 9.0 |  |
| **MOS social support scale** |  |  |  | 0.781 |
| Sample n | 54 | 45 | 99 |  |
| Median | 4.8 | 4.9 | 4.8 |  |
| Q1, Q3 | 4.3, 5.0 | 3.9, 5.0 | 4.2, 5.0 |  |
| Range | 2.9 - 5.0 | 2.0 - 5.0 | 2.0 - 5.0 |  |

##### Validity check

###### HSCL scores of distress showed good agreement with the SRQ-20

The Bland-Altman plot of mean score on the HSCL and the rescaled total score on the SRQ-20 (Figure S2) showed a reasonably even scatter of dots, with most dots lying within the 95% confidence interval of the difference between measurements. The mean difference between measurements of slightly less than 0.2 suggests that, on average, rescaled SRQ-20 ratings exceed mean HSCL ratings by about 0.2 points, or about 7% of the range (i.e. a bias towards slightly higher estimates from the SRQ-20 than from the HSCL). Three (of 99) observations lay above, and two lay below, the 95% CI. The estimated slope of the regression line was 1.00, with 95% confidence interval bounds of 0.88 and 1.13 (Figure S2). Lin’s concordance correlation coefficient (95% CI) was 0.81 (0.74 - 0.87) . Together, these results indicate that the estimates of distress yielded by the HSCL tool show good agreement with the estimates of distress yielded by the SRQ-20.

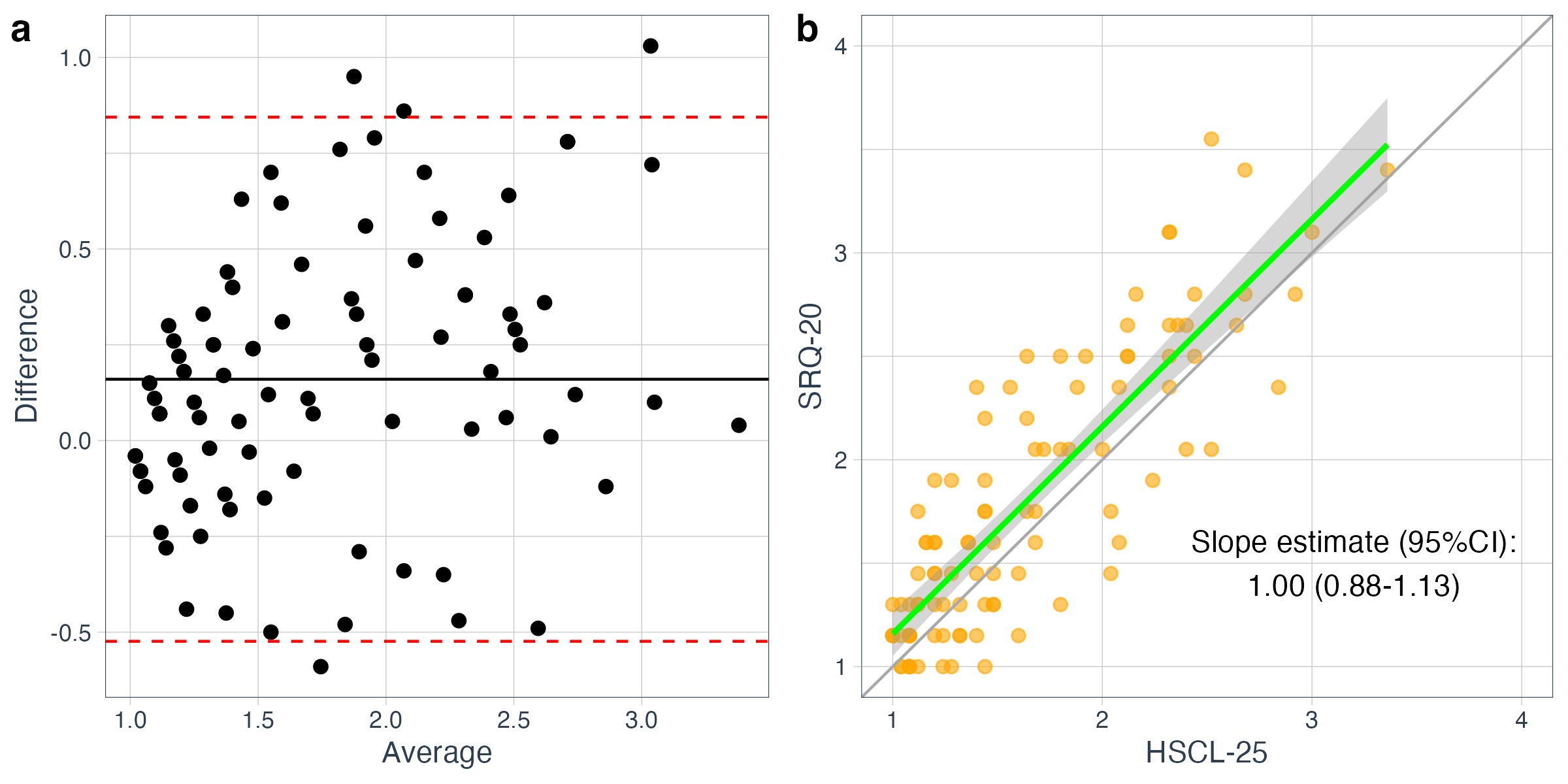

Figure S2: Assessment of agreement between HSCL and SRQ-20 scores, using (a) Bland-Altman plot and (b) regression with Lin’s concordance correlation coefficient, showed good agreement.

#### Missing ratings of HFS trains

Ratings were missing for 119 of 225 HFS trains delivered, due to technical error. Inspection of the existing data showed that ratings increased systematically with each train (Figure S3a) and were non-parametrically distributed within each train (Figure S3a, Table S3). Therefore, the missing ratings were imputed using the median of the available ratings for the relevant train. The within-participant median of the 5 trains, including the imputed data, was used as the adjustment variable for the models used to test Hypothesis 2.

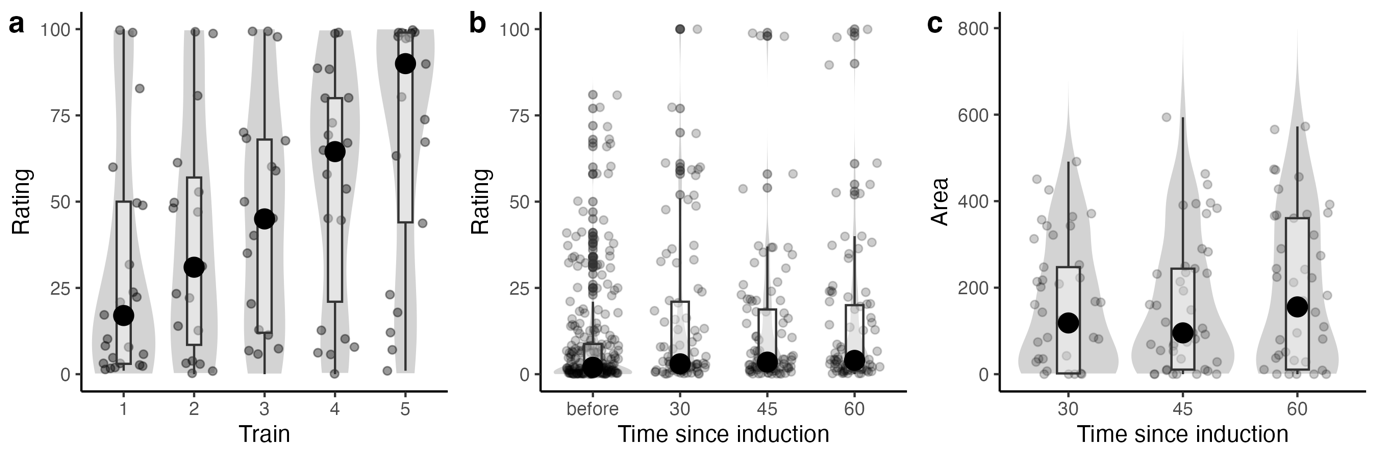

Figure S3: (a) VAS ratings increased with successive trains of high-frequency electrical stimulation. (b) VAS ratings of pinprick stimuli were generally higher after induction than before. (c) The sample-average surface area of induced SH exceeded 0 at all 3 assessment times.

Table S3: Pinprick ratings and SH outcomes by study group.

|  | pain-free (N=26) | persist-pain (N=19) | Total (N=45) | p value |
| --- | --- | --- | --- | --- |
| **Mean rating before induction** |  |  |  | 0.046 |
| Median | 1.0 | 7.5 | 1.7 |  |
| Q1, Q3 | 0.3, 3.4 | 0.7, 22.7 | 0.3, 8.8 |  |
| Range | 0.0 - 57.0 | 0.0 - 58.7 | 0.0 - 58.7 |  |
| **Mean rating 30 mins after induction** |  |  |  | 0.263 |
| Median | 2.0 | 16.0 | 3.5 |  |
| Q1, Q3 | 1.5, 9.6 | 1.2, 29.8 | 1.5, 24.0 |  |
| Range | 0.0 - 88.5 | 0.0 - 80.0 | 0.0 - 88.5 |  |
| **Mean rating 45 mins after induction** |  |  |  | 0.115 |
| Median | 2.5 | 11.5 | 3.5 |  |
| Q1, Q3 | 1.2, 14.8 | 1.8, 27.0 | 1.5, 18.0 |  |
| Range | 0.0 - 98.0 | 0.0 - 68.0 | 0.0 - 98.0 |  |
| **Mean rating 60 mins after induction** |  |  |  | 0.131 |
| Median | 3.0 | 13.0 | 3.5 |  |
| Q1, Q3 | 1.1, 13.9 | 1.5, 29.8 | 1.5, 18.0 |  |
| Range | 0.0 - 94.5 | 0.0 - 99.0 | 0.0 - 99.0 |  |
| **Surface area of SH 30 mins after induction** |  |  |  | 0.124 |
| Median | 77.8 | 213.9 | 118.4 |  |
| Q1, Q3 | 0.4, 193.1 | 73.4, 317.3 | 1.8, 247.5 |  |
| Range | 0.0 - 426.0 | 0.0 - 491.4 | 0.0 - 491.4 |  |
| **Surface area of SH 45 mins after induction** |  |  |  | 0.193 |
| Median | 72.5 | 148.5 | 95.5 |  |
| Q1, Q3 | 9.3, 208.6 | 52.1, 263.4 | 10.6, 244.0 |  |
| Range | 0.0 - 463.2 | 0.0 - 594.0 | 0.0 - 594.0 |  |
| **Surface area of SH 60 mins after induction** |  |  |  | 0.298 |
| Median | 102.5 | 244.0 | 155.6 |  |
| Q1, Q3 | 15.9, 237.8 | 18.6, 368.6 | 10.6, 360.6 |  |
| Range | 0.0 - 572.8 | 0.0 - 565.7 | 0.0 - 572.8 |  |
| **Magnitude of SH 30 mins after induction** |  |  |  | 0.462 |
| Median | 1.4 | 0.5 | 1.3 |  |
| Q1, Q3 | 0.0, 8.9 | -0.1, 13.4 | 0.0, 10.2 |  |
| Range | -2.7 - 37.7 | -9.3 - 56.8 | -9.3 - 56.8 |  |
| **Magnitude of SH 45 mins after induction** |  |  |  | 0.774 |
| Median | 1.2 | 0.8 | 1.0 |  |
| Q1, Q3 | 0.0, 3.8 | -1.7, 8.0 | 0.0, 6.7 |  |
| Range | -2.7 - 41.0 | -12.0 - 24.8 | -12.0 - 41.0 |  |
| **Magnitude of SH 60 mins after induction** |  |  |  | 0.927 |
| Median | 1.0 | 0.8 | 0.8 |  |
| Q1, Q3 | 0.0, 4.2 | -0.7, 12.2 | 0.0, 9.2 |  |
| Range | -3.3 - 37.5 | -16.7 - 40.3 | -16.7 - 40.3 |  |

#### Exploratory analysis

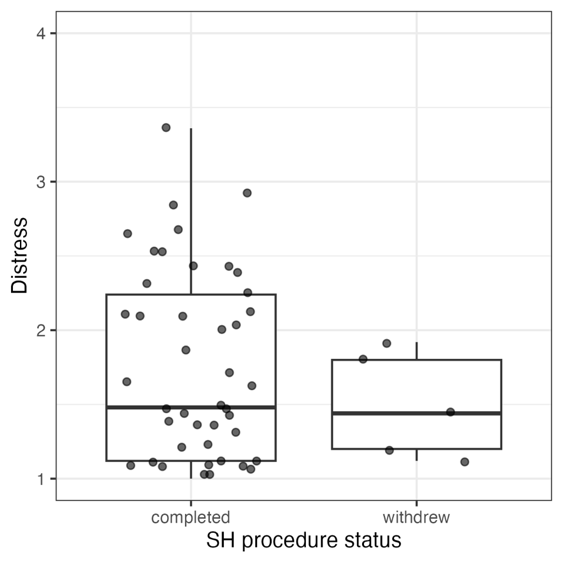

Figure S4: Distress ratings were not appreciably different between participants who completed and participants who withdrew from the SH procedure.

#### References used in supplementary file

[1] Blough DS. A method for obtaining psychophysical thresholds from the pigeon. J Exp Anal Behav 1958;1(1):31-43.

[2] Goodrich B, Gabry J, Ali I, Brilleman S. rstanarm: Bayesian applied regression modeling via Stan, 2024.
